# Oropouche Virus Outbreak in Southeast, Brazil: Expanding Beyond the Amazonian Endemic Region

**DOI:** 10.1101/2024.12.11.24318883

**Authors:** Edson Delatorre, Gabriela Mendonça de Colombo, Felipe Donateli Gatti, Anna Clara Gregório Có, Julia del Piero Pereira, Eric Arrivabene Tavares, Joana Zorzal Nodari, Agata Rossi, Suwellen Sardinha Dias de Azevedo, Cláudio Tavares Sacchi, Karoline Rodrigues Campos, Adriana Bugno, Lyvia Neves Rebello Alves, Lucas André Silva Bonela, Jaqueline Pegoretti Goulart, Thiago de Jesus Souza, Felipe Gomes Naveca, Rodrigo Ribeiro-Rodrigues

## Abstract

The Oropouche virus (OROV), historically endemic to the Amazon, has spread to nearly all Brazilian states in 2024, with Espírito Santo emerging as a significant transmission hotspot in the Atlantic Forest biome. We characterized the epidemiological factors driving OROV spread in non-endemic Southeast Brazil, analyzing environmental and agricultural conditions contributing to viral transmission. Samples from 29,000 suspected arbovirus-infected patients were tested by RT-qPCR for Dengue, Chikungunya, Zika, Mayaro, and Oropouche virus. Between March and June 2024, OROV cases in the state reached 339, demonstrating successful local transmission.

Spatial analysis revealed that most cases clustered in municipalities with tropical climates and intensive cacao, robusta coffee, coconut, and pepper cultivation. Phylogenetic analysis identified the Espírito Santo OROV strains as part of the 2022-2024 Amazonian lineage. The rapid spread of OROV outside the Amazon highlights its adaptive potential and public health threat, emphasizing the need for enhanced surveillance and targeted control measures.

## Introduction

The Oropouche virus (OROV), classified into the *Orthobunyavirus oropoucheense* species within the family *Peribunyaviridae*, is an arthropod-borne virus with a negative-sense RNA genome consisting of three segments: Large (L), Medium (M), and Small (S) ^1^. This neglected arbovirus causes Oropouche fever and circulates primarily in Central America, South America, and the Caribbean ^2–4^. In Brazil, OROV was historically confined to the Amazon basin, where a range of reservoirs maintain its sylvatic transmission cycle, facilitated by several vector species ^2,3,5^. In Amazonian urban areas, *Culicoides paraensis*, a midge commonly found in tropical, humid environments rich in organic matter — such as forests and plantations — is the primary vector responsible for OROV transmission to humans ^3,4^. Humans may acquire OROV infection in forested regions and subsequently introduce it to urban settings. The widespread distribution of this vector, coupled with increased human mobility and the influence of climate change, may facilitate the virus’s gradual expansion beyond its historical range in Brazil, raising concerns about the potential for broader geographic spread ^6–8^.

Since the 1960s, occasional OROV spillovers to humans have led to at least 30 documented localized outbreaks or large-scale epidemics in the Amazon basin, underscoring the virus’s epidemic potential ^9–17^. Although its incidence is most prominent in the Amazon region, sporadic detection has also been documented in other Brazilian states, such as Bahia, Goiás, Maranhão, Mato Grosso, Mato Grosso do Sul, and Minas Gerais. However, these occurrences have not led to widespread human outbreaks^18^. Between August 2022 and March 2024, a new outbreak triggered by a reassortant OROV lineage emerged in Brazil’s western Amazon region, resulting in nearly 6,000 reported cases^19^. In 2024, this reassortant lineage triggered the largest recorded outbreak outside the virus’s endemic zone, with OROV detected in all Brazilian regions^20,21^. Outside the Amazon, high incidence rates were observed in the Atlantic Forest region, particularly in municipalities with low population densities and agricultural activities favoring the establishment of *C. paraensis* vector populations, such as cocoa and banana cultivation^22^.

Significant changes in disease manifestation and severity have accompanied the unprecedented geographic expansion of this new reassortant OROV lineage. OROV infections typically present as an acute febrile illness characterized by headache, myalgia, and arthralgia^19^. These symptoms overlap with those of other endemic arboviruses, such as dengue (DENV), Zika (ZIKV), and chikungunya (CHIKV)^2,3^. However, emerging evidence has linked the virus to fatal cases for the first time ^23^. Unprecedented vertical transmission was also reported, and some cases resulted in congenital anomalies or fetal death^24,25^. Therefore, this significant shift in OROV’s pathogenicity marks a new epidemiological paradigm for Oropouche fever.

Espírito Santo state, located in southeastern Brazil and entirely within the Atlantic Forest biome, has emerged as a significant hotspot for OROV transmission outside the Amazon region, recording the highest incidence rate among non-Amazonian states in 2024^26^. The state’s extensive agricultural activities, particularly in coffee, cocoa, and banana cultivation^27^, combined with high rural worker mobility and environmental conditions favorable for *C. paraensis* establishment, may have contributed to viral spread. Here, we employ epidemiological and genomic approaches to analyze the emergence and dissemination of the new OROV variant in Espírito Santo state. We examine the regional characteristics that facilitate its transmission and contribute to its establishment in this previously unaffected area.

## Materials and Methods

### Study Population

This research was conducted using samples from patients who visited public health units in Espírito Santo state with symptoms suggestive of arboviral infection (ZDC: Zika, dengue, and chikungunya) between February 25 and June 15, 2024. The serum or plasma samples were subsequently sent to the LACEN-ES, Brazil, for viral molecular diagnosis. This study was approved by the Ethics Committee of the University of Vila Velha, Espírito Santo - Brazil (CAAE: 84698324.7.0000.5064). The Committee waived the requirement for written informed consent.

### Sample processing and Real-time RT-PCR

Samples were processed for nucleic acid extraction using magnetic bead-based systems, including the TANBead Maelstrom™ 9600, Loccus EXTRACTA® 96, and TechStar YC-702. The extraction was performed according to the manufacturers’ instructions, utilizing kits specifically compatible with each system: the Extracta MDx DNA and RNA Pathogen Kit (Loccus) and the Magnetic Bead-based DNA/RNA Viral Nucleic Acid Extraction Kit (TechStar Technology). Subsequently, for molecular testing, three different Reverse Transcription Polymerase Chain Reaction (RT-qPCR) kits were employed at different intervals throughout the study period: MOLECULAR ZCD TIPAGEM Bio-Manguinhos, BIOMOL ZDC IBMP, and VIASURE Zika, Dengue & Chikungunya, according to the protocols provided by the manufacturers.

For the samples not detectable for ZCD, a multiplex RT-qPCR was carried out to investigate for Oropouche and Mayaro viruses^28^. As a way of extending testing, the laboratory diagnosis was adapted to carry out the procedure on a pool of samples. Each pool was made up of eight samples, and the volumes added followed the proportion used in the original protocol. After detecting the target, another RT-qPCR was conducted with the eight samples individually, and the test was confirmed.

### Epidemiological data and environmental context

Individual-level of OROV-positive case data were retrieved from the *eSUS* (https://sisaps.saude.gov.br/esus/) and *Gerenciador de Ambiente Laboratorial* (http://gal.datasus.gov.br/) systems at the municipal level for the state of Espírito Santo, covering cases reported up to June 2024. Each case was associated with a notification code and metadata, which included demographic information, spatial location, laboratory results, and clinical details such as symptoms, hospitalization status, and dates related to notification and symptom onset, with this information available for most cases. All identifiers were anonymized.

The municipal incidence of OROV was calculated based on the 2022 census data from the Brazilian Institute of Geography and Statistics (IBGE, https://www.ibge.gov.br/en/statistics/social/labor/22836-2022-census-3.html). The mapping of the estimated incidence values used the “geoBR” package in RStudio, based on municipal boundaries shapefiles from IBGE (https://www.ibge.gov.br/en/geosciences/downloads-geosciences.html). Data about the economic activities conducted in agricultural establishments at the municipal level were obtained from the 2017 census of agriculture conducted by IBGE (https://censoagro2017.ibge.gov.br). We estimated the Spearman correlation for all municipalities reporting cases to explore the relationship between the planted area of the top ten crops in Espírito Santo and the number of OROV cases.

### Generation Time and Instantaneous Reproduction Number Estimation

The generation time represents the interval between successive rounds of infection, precisely the time between the infection of a primary human case and the subsequent infection of secondary human cases caused by the primary case. Although the generation time for other arboviruses has been estimated, currently, there are no specific estimates for OROV. We estimated the generation time using a combination of human viral clearance data^29^, mosquito mortality rates^30^, and data from experimental studies involving vector species^31^, using a framework applied previously for ZIKV^32^ and MAYV^33^. Parameter inference for the generation time was conducted using a Bayesian framework implemented with Markov chain Monte Carlo (MCMC) methods. The posterior distributions of the generation time parameters were then used to inform the calculation of the reproduction number. We estimated the instantaneous reproduction number, Rt, for OROV using the R package EpiEstim^34^. The Rt was reconstructed for the April-June period using a Bayesian inference model with a sliding time window. Further technical details regarding the MCMC implementation, likelihood functions, and priors can be found in Supplementary Material (Methods).

### Sample selection and Next Generation Sequencing

Seven representative samples with Ct < 27 from the most affected municipalities were selected for complete genome sequencing. Six samples were sequenced at Instituto Adolfo Lutz (IAL), resulting in complete S and M segments (Supplementary Material, Table S1). One additional sample was sequenced at LACEN/ES using the Illumina RNA Prep with Enrichment (L) Tagmentation protocol with the Respiratory Pathogen ID/AMR Enrichment Panel (RPIP) Kit (Illumina 2021). OROV reads were extracted from the non-target portion of the RPIP kit. Genome assembly was conducted using a custom version of the ViralFlow pipeline v1.0.1^35^, referencing NCBI sequences NC_005776.1 (L segment), NC_005775.1 (M segment), and NC_005777.1 (S segment).

### Bioinformatics and Phylogenetic Inference

For the phylogenetic analysis, we aligned the seven newly generated sequences from ES with the complete genome of 145 OROV strains sampled in the Americas between 1955 and 2023 and available at GenBank until August 2024. The alignment was conducted in MAFFT 7 using the L-INS-i strategy^36^. It included the prototypes of OROV, Iquitos virus (IQTV), Madre de Dios virus (MDDV), and Perdões virus (PEDV), as outgroups. The GTR+G substitution model was selected for all segments by the Corrected Akaike Information Criterion in jModelTest2^37^. The phylogenetic tree was inferred using Bayesian inference in MrBayes 3.2.7a^38^ with eight simultaneous Markov chain Monte Carlo (MCMC) iterations of 2,700,000, 11,300,000, and 3,900,000 generations for the S, M, and L genome segments, respectively. Trees were sampled every 1,000 generations, with a burn-in of 25%, until the convergence diagnostic met the stopping rule (standard deviation of split frequencies < 0.01), satisfactory Estimated Sample Size (ESS > 200), and Potential Scale Reduction Factor (PSRF) approaching 1.0. Node support was determined by posterior probabilities (*PP*) directly estimated from the majority rule consensus topology.

## Results

### Rapid Emergence of OROV Amid Arbovirus Circulation in Espírito Santo with Shifting Epidemiological Trends

Approximately 29,100 patients in Espírito Santo presenting arbovirus-like illness were tested by real-time RT-PCR for the presence of DENV-1, DENV-2, CHIKV, ZIKV, OROV, and MAYV from March to June 2024 (Figure 1). Until epidemiological week (EW) 13, the arbovirus circulation was dominated by DENV-1, representing approximately 50% of the positive cases, followed by CHIKV and DENV-2 (Figure 1A). However, from EW 13 onwards, the first cases of OROV were detected, marking a pronounced shift in the epidemiological landscape. Over the subsequent weeks, OROV cases increased rapidly, reaching 339 cases within 10 weeks. By EW 24, the frequency of OROV infections approached the levels of DENV-1, DENV-2, and CHIKV, indicating a comparable circulation of these viruses at the outbreak’s peak. Following the emergence of OROV, the proportion of CHIKV cases also rose, eventually surpassing that of DENV. Initially, OROV cases were primarily classified as imported; however, community spread became evident as local transmission was established (Figure 1B). The transmissibility of OROV in Espírito Santo was estimated based on the time-varying reproduction number (Rt). Between EW 18 and 27, the estimated Rt for OROV remained approximately 2.5, with a peak value of ∼3.0 observed in EW 26. Following this peak, a marked reduction in Rt occurred, falling below 1.0 by EW 28, indicating a decline in transmission and a shift towards containment of the outbreak (Figure 1B).

**Figure 1.**
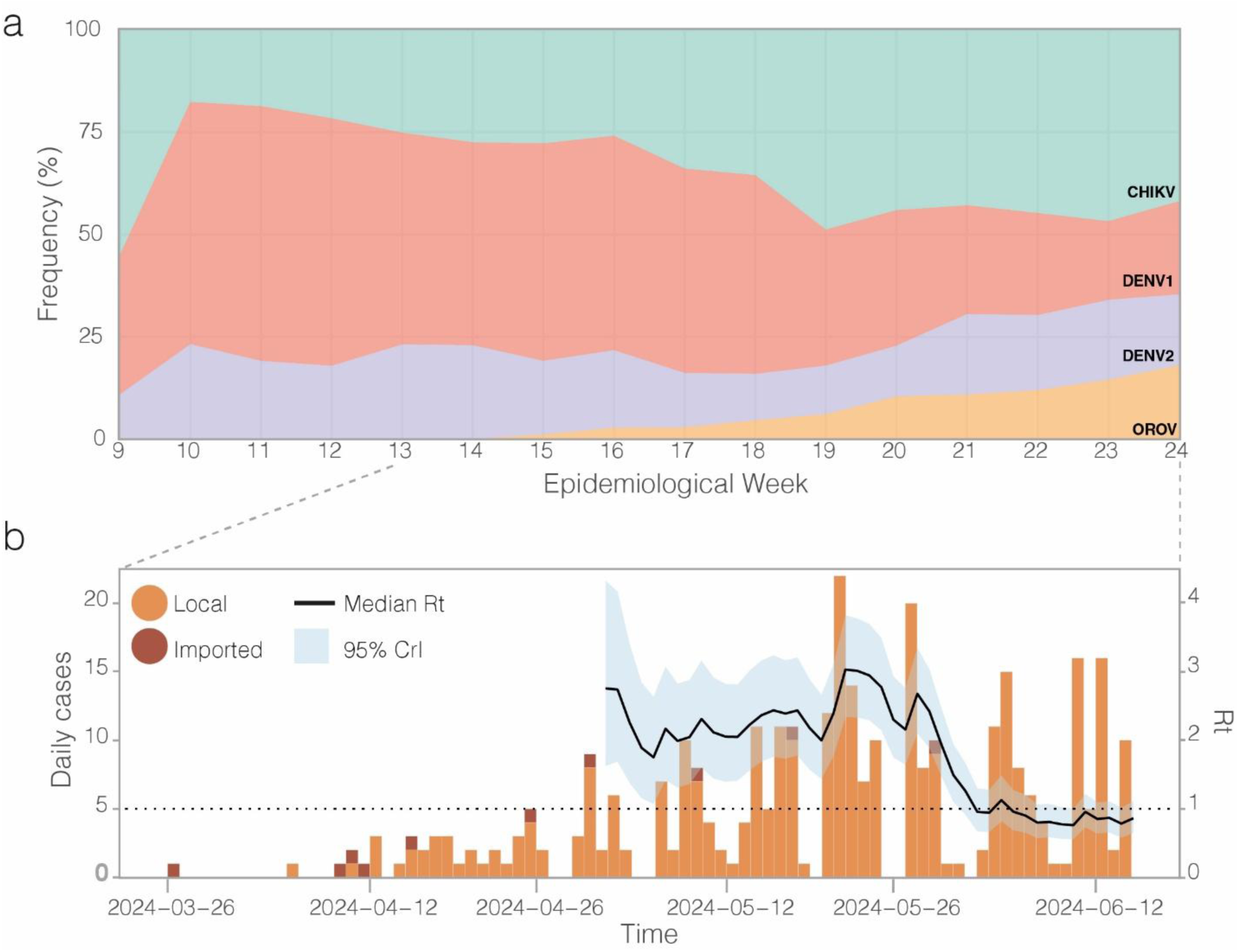
Temporal Distribution and Reproduction Number of Oropouche Virus (OROV) in Espírito Santo. a) Weekly proportions of chikungunya virus (CHIKV), Dengue virus type 1 (DENV-1), type 2 (DENV-2), and OROV in Espírito Santo, from epidemiological weeks 9 to 24, 2024. b) Estimates of the instantaneous reproduction number (Rt) during the OROV outbreak (mean (solid line) and 95% credible interval (shaded area)) based on daily incidence data covering local and imported cases. The reproduction number was estimated using sliding windows of τ = 6 days.

### Oropouche Outbreak in Espírito Santo predominates in working-age men and shows arboviral-like symptoms

The demographic analysis of OROV cases revealed a marked predominance of male patients (Figure 2A), with a male-to-female ratio of 1.5:1. Most cases occurred in individuals older than 20 years, suggesting that adult males may be disproportionately affected by this outbreak. Fever (90.56%), headache (81.12%), myalgia (68.44%), and retro-orbital pain (33.33%) were the most frequently reported symptoms (Figure 2B). Notably, no significant differences in symptom presentation were observed between male and female patients. Real-time RT-PCR Ct values were employed as a proxy for the patient’s plasmatic viral load to investigate its relationship and symptomatology. Patients with fever exhibited significantly lower median Ct values compared to those without fever (Mann–Whitney test, *P* = 0.017), suggesting a higher viral load in febrile individuals. A similar trend was observed for headache (*P* = 0.043), myalgia (*P* = 0.025), and retro-orbital pain (*P* = 0.017) symptoms. Comparisons for other symptoms did not yield statistically significant differences (Suplementary Material, Figure S1).

**Figure 2.**
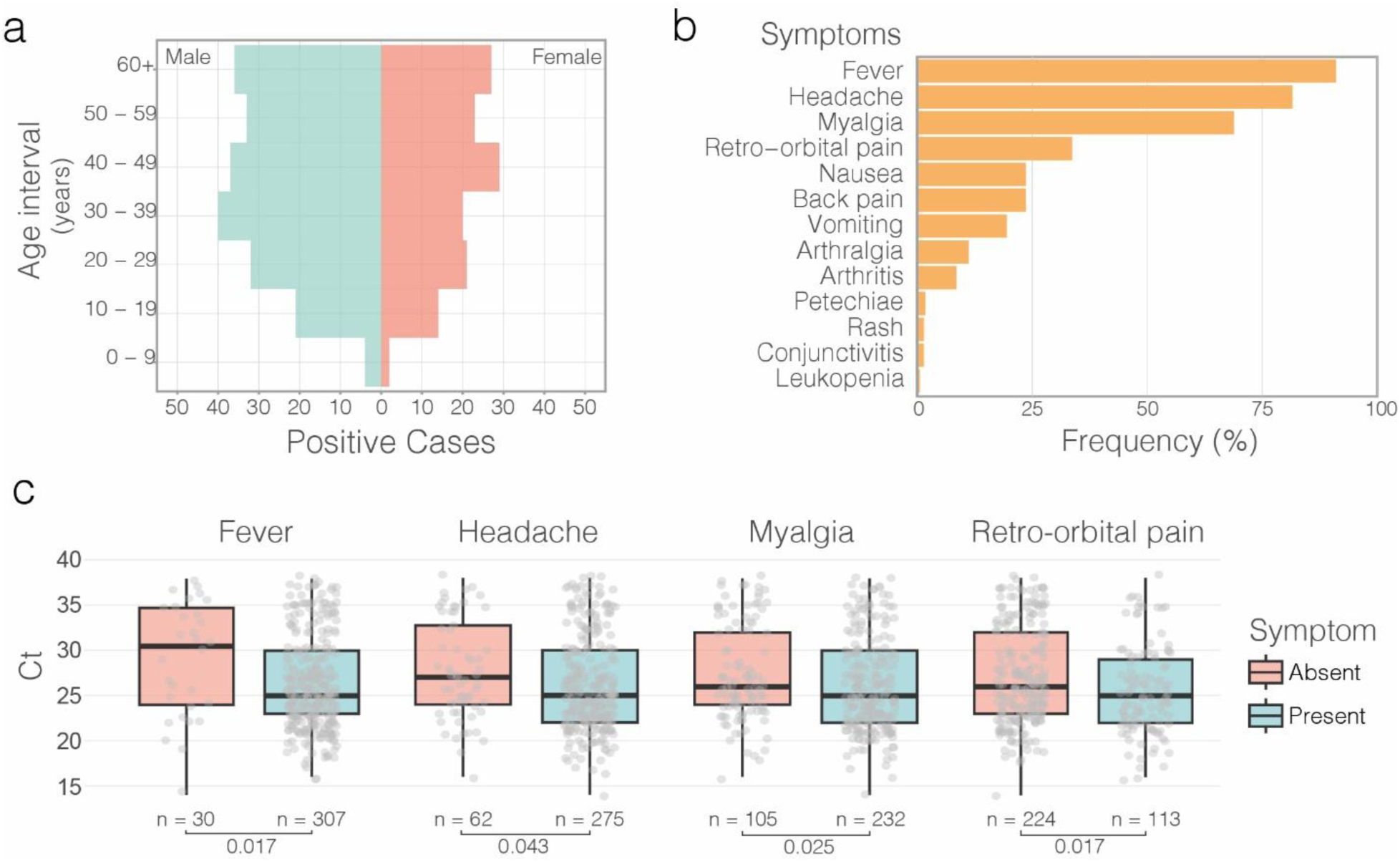
Demographics, Symptoms, and Ct Value Comparison in OROV-positive Patients from Espírito Santo. a) Age-sex pyramid of the OROV-positive population from Espírito Santo, with males on the left and females on the right. b) Frequency of symptoms observed in OROV-positive patients. c) Comparison of Ct values from OROV RT-PCR in patients presenting or not presenting symptoms of fever, headache, myalgia, and retro-orbital pain. The boxplots compare the Ct values of individuals with and without each symptom. Horizontal bars represent the Ct medians and interquartile ranges (IQR). Two-sided P-values for the nonparametric Mann–Whitney test are shown for each group.

**Figure 3.**
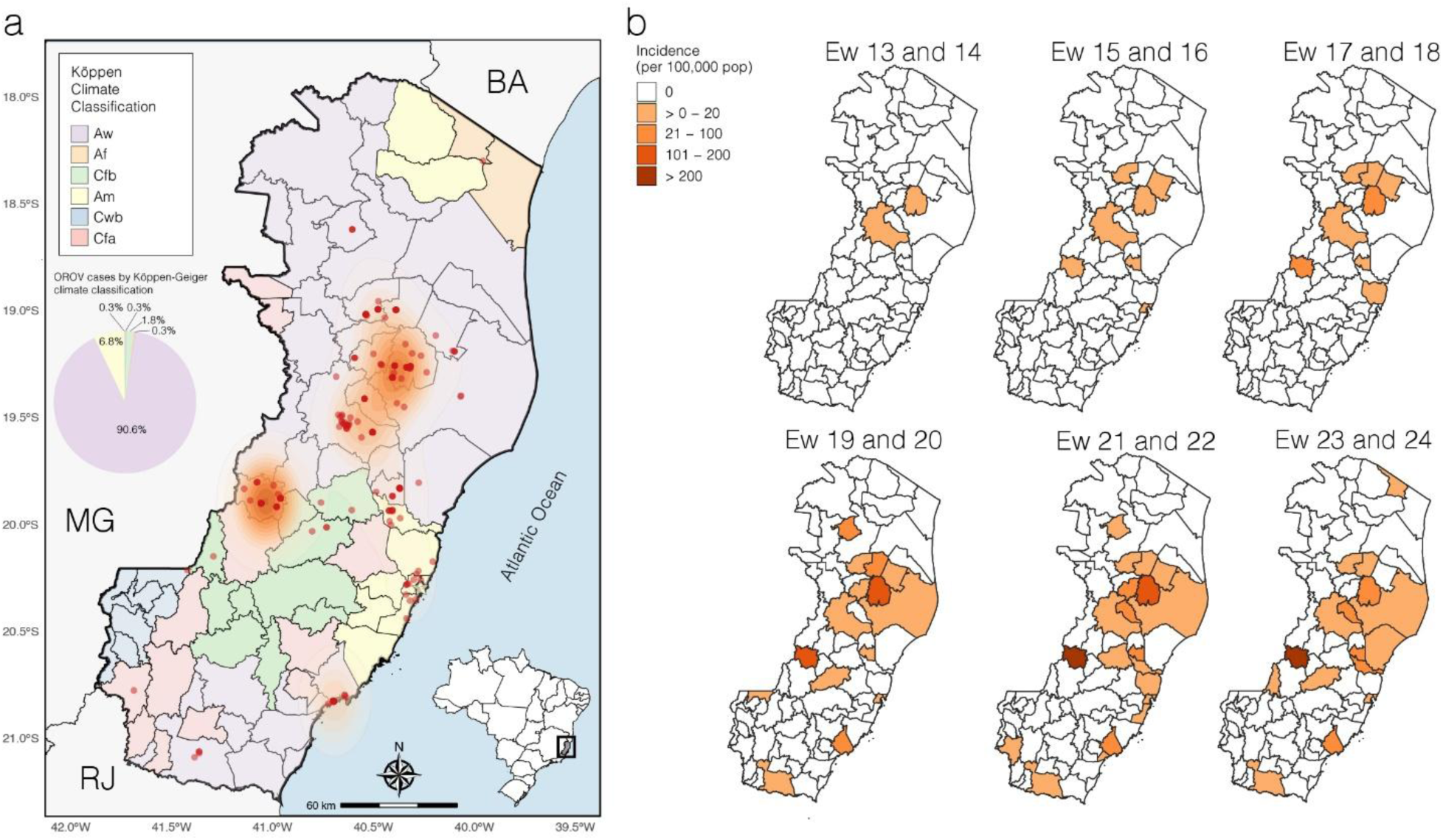
Spatial Analysis of OROV Cases and Climate Classification in Espírito Santo. a) Map of Espírito Santo, with municipalities colored according to the Köppen climate classification (legend included). Red points represent OROV cases, and an orange shading depicts case density. The pie chart illustrates the proportion of cases in each Köppen climate. b) Incidence of OROV cases in each municipality, represented by the color gradient, calculated for each two-week epidemiological period.

### Spatial-Temporal Spread of OROV Was Associated with Tropical Climates in Two Main Hotspots

During the OROV outbreak in Espírito Santo, the virus spread across 17 municipalities, culminating in 8.84 cases per 100,000 inhabitants statewide. However, most cases were concentrated in two distinct hotspots: the regions surrounding the municipalities of Colatina/Rio Bananal, and Laranja da Terra. The spatial distribution of OROV cases revealed a significant association with specific climatic profiles. Remarkably, 98% of the diagnosed cases were associated with tropical climates, with approximately 91% occurring in municipalities classified as having a tropical savanna climate (Köppen Aw), and an additional 7% linked to regions characterized by a tropical monsoon climate (Köppen Am). In contrast, only ∼2% of cases were distributed across municipalities with temperate climates, such as humid subtropical (Köppen Cfa) and temperate oceanic (Köppen Cfb). Notably, no cases were reported in areas classified under the subtropical highland climate (Köppen Cwb). These findings highlight potential environmental or ecological factors limiting transmission in these regions.

The initial spread of OROV in Espírito Santo followed a clear spatial pattern, predominantly affecting municipalities within the tropical savanna climate. This dissemination phase, between EW 17 and 18, marked the entry of the virus into areas with different climatic conditions. The peak incidence, exceeding 200 cases per 100,000 inhabitants, was recorded between EW 21 and 24, particularly in the epicentral municipalities of Colatina and Laranja da Terra. These areas, situated within the Aw climate zone, appear to have served as primary foci for transmitting OROV throughout the state.

### Oropouche prevalence is associated with Cocoa, Robusta Coffee, Coconut, and Pepper plantations

To further characterize the ecological niches contributing to the introduction and spread of OROV in Espírito Santo, we explored the association between OROV prevalence and the cultivated areas for the top ten crops in the state, as reported by the IBGE agricultural census. The Spearman correlation test showed significant associations between specific crops and OROV incidence (Figure 4). Notably, robusta coffee (ρ = 0.55, *P* = 0.0044), cacao (ρ = 0.54, *P* = 0.0054), coconut (ρ = 0.43, *P* = 0.00301), and pepper (ρ = 0.43, *P* = 0.0339) displayed moderate positive correlations with the number of OROV cases. These results indicate that agricultural practices, especially in areas dominated by these crops, may have influenced the local dynamics of OROV transmission, possibly by affecting mosquito habitats or increasing human exposure.

**Figure 4.**
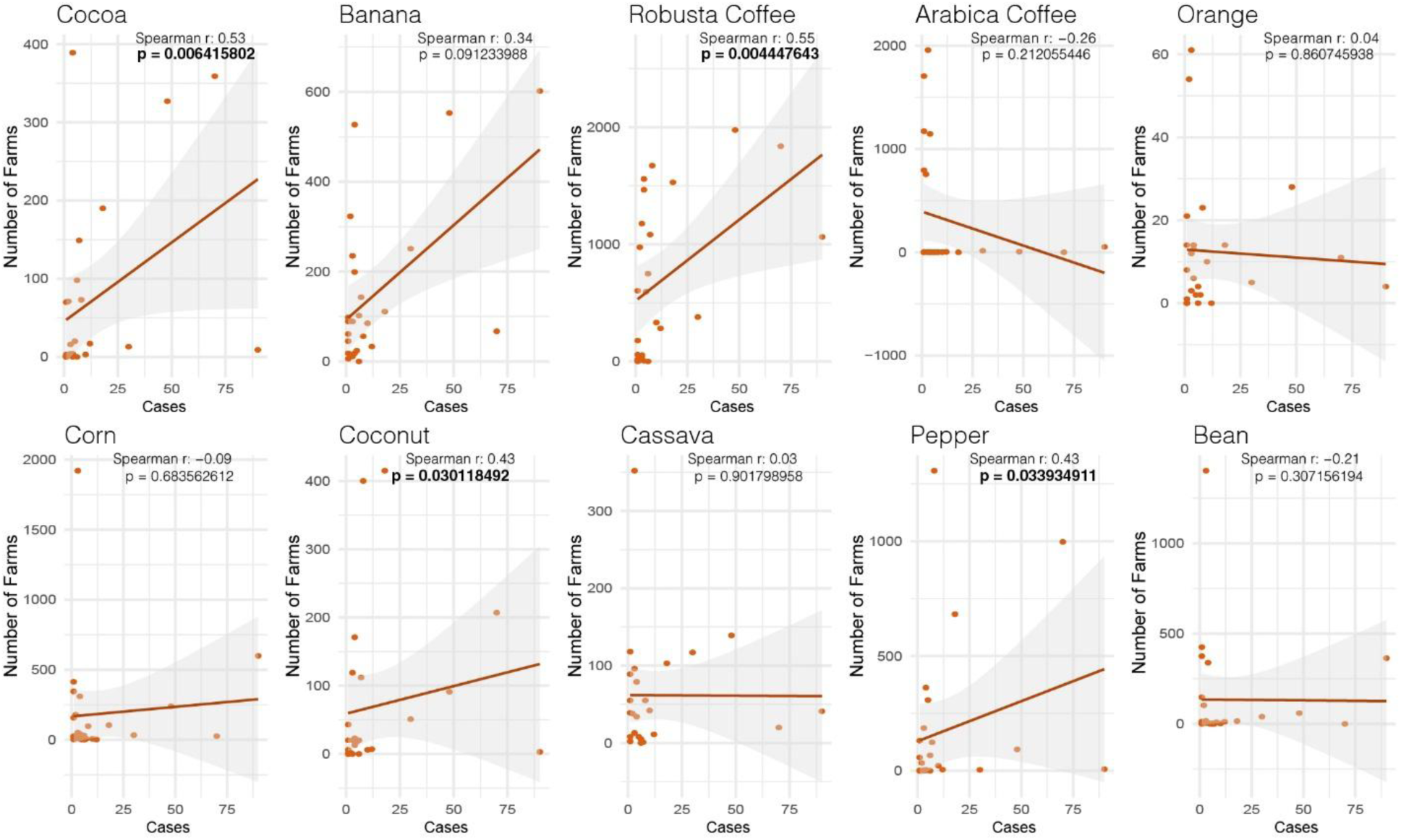
Correlation Between Agricultural Activity and OROV Case Counts. Series of correlation plots demonstrating the relationship between the number of farms cultivating each of the top 10 crops in Espírito Santo and the number of confirmed OROV cases per municipality. Each plot shows the non-parametric Spearman correlation coefficient (rho), along with 95% confidence intervals and p-values, providing insight into each association’s strength, direction, and statistical significance. Statistically significant p-values are highlighted in bold.

### OROV Strains from Espírito Santo are linked with the 2022-2024 Amazonian Outbreak

Bayesian inference of the OROV genome sequences from Espírito Santo revealed that these samples cluster within the Brazil 2015–2024 clade, alongside samples from the Amazon region (Figure 5). Phylogenetic analysis of each genomic segment individually showed that all cases detected outside Espírito Santo belong to the novel reassortant M1L2S2 (OROV_BR2025-2024_) lineage^19^ with high branch support (*PP* ≥ 0.98), which circulated in the Amazon Basin between 2023 and 2024, causing a major outbreak. Interestingly, in the S and M trees, the ES samples do not form a monophyletic clade, indicating multiple introductions of OROV into the state (Figure 5A and B). This suggests that the spread of OROV into Espírito Santo followed a complex pattern, with multiple viral introductions.

**Figure 5.**
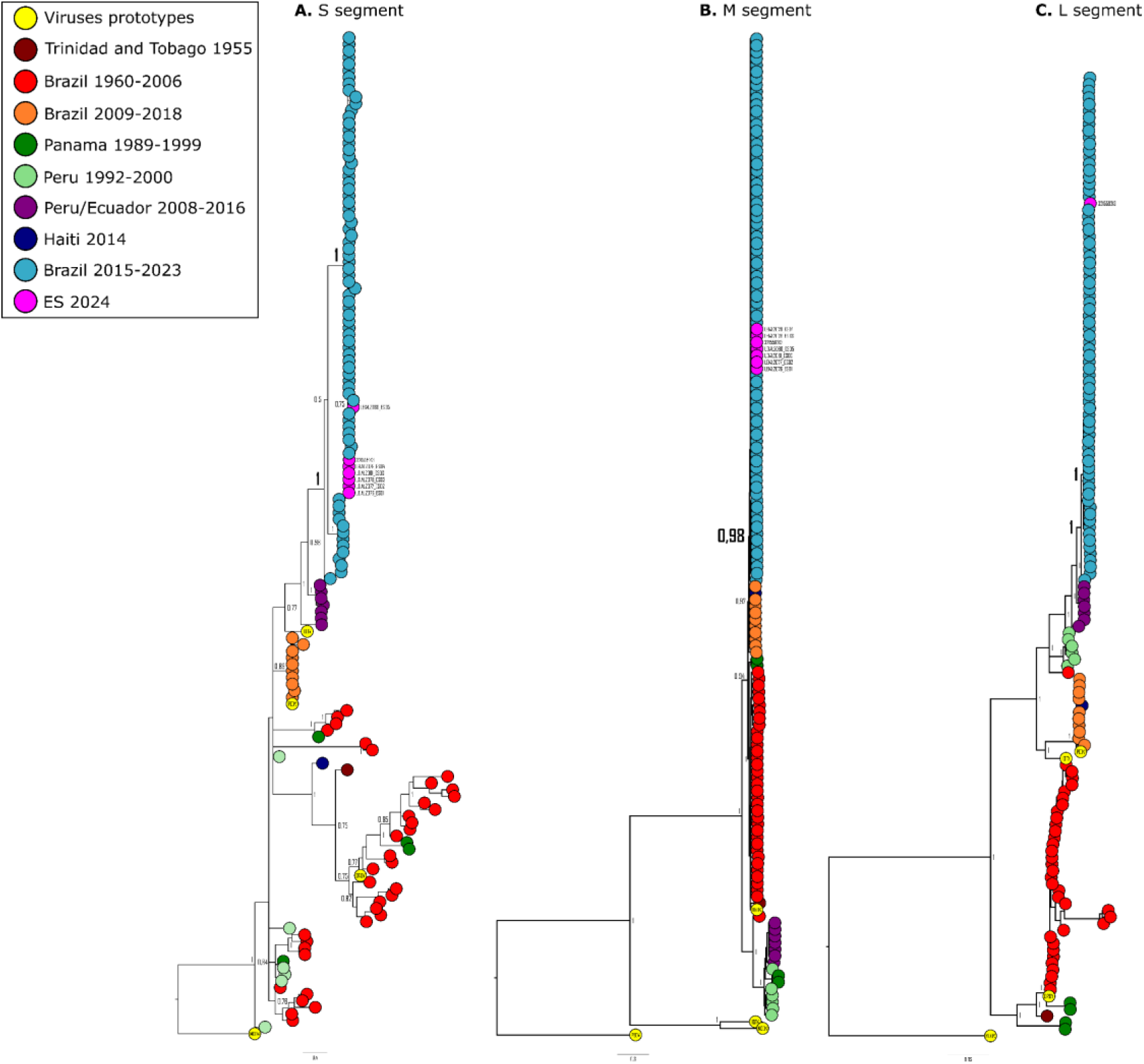
Phylogenetic Analysis of OROV Segments L, M, and. **S.** Bayesian phylogenetic trees of OROV segments S (a), M (b), and L (c), associated with the reassortant lineage M1L2S2. Each tree includes a representative subsample of genomes from the epidemic clade (highlighted), with tip colors indicating sampling locations. Branch support values are provided for the main clades.

## Discussion

The emergence and rapid spread of arboviruses beyond their traditional endemic regions presents a growing global health challenge, intensified by changing climate patterns and human land use^8^. Our study documents, using high-resolution data, the unprecedented establishment of OROV in Brazil’s Atlantic Forest biome, demonstrating how a pathogen historically confined to the Amazon Basil can adapt and spread within new ecological settings. This expansion mirrors patterns observed for other arboviruses, such as Zika^39^, Chikungunya^40^, and West Nile virus^41^, where changes in vector ecology, human mobility, and environmental conditions have facilitated the emergence of these viruses in previously unaffected regions^7^. OROV’s successful establishment in Southeast Brazil underscores both an immediate public health concern and the complex ecological and epidemiological factors enabling arboviral adaptation to novel environments.

Our findings reveal a marked epidemiological shift with the emergence and rapid establishment of OROV transmission in the state amidst the ongoing endemic circulation of DENV and CHIKV. Within approximately 11 weeks from its initial detection, OROV reached infection levels comparable to those of endemic arboviruses, suggesting robust viral transmission potentially facilitated by a largely susceptible population and high densities of vectors. The concurrent presence of OROV alongside DENV and CHIKV, without evident competitive interference, could reflect vector-specific ecological differences: while DENV and CHIKV predominantly utilize *Aedes* mosquitoes in urban areas^42^, OROV transmission is driven mainly by *C. paraensis* midges, which thrive in rural and periurban environments^3,29^. The estimated transmissibility, with an Rt peaking at approximately 3.0, parallels the dynamics seen in both urban and sylvatic arboviruses within immunologically naive or mixed populations. Similar patterns were reported for ZIKV’s spread, with Rt values ranging from 2.6 to 4.8 in French Polynesia^43,44^, 3.0 to 6.6 in Colombia ^45^,and reaching values of 3.6 and 2.5 in Singapore^46^ and Brazil^32^, respectively. Likewise, the Mayaro virus, a sylvatic arbovirus endemic to the Amazon, exhibits Rt values of 2.1-2.9 within its primary ecological range, although its transmissibility (Rt: 1.1-1.3) declines in non-Amazonian regions^33^. The high transmissibility of OROV observed in Espírito Santo could be attributed to multiple factors, including the predominantly mild or asymptomatic nature of OROV infections^3,29^ allowing infected individuals to remain mobile and maintain transmission chains and favorable local conditions for *C. paraensis* proliferation^30^.

The demographic profile of affected individuals in this outbreak also reflects unique eco-epidemiological features, with a predominance of adult male cases that contrasts with previous OROV outbreaks showing balanced or slightly female-skewed distributions^16,19,21,47^. This male predominance may reflect occupational exposure patterns in rural settings, where activities associated with agriculture elevate contact with *C. paraensis* vectors, consistent with past Brazilian outbreaks in which farming occupations represented a significant risk factor^47^. Clinical manifestations align with previous OROV cases across diverse regions, including recent Amazonian outbreaks^16,19,48^. The association between key symptoms and lower Ct values suggests that higher viral loads may drive the intensity of clinical manifestations. This relationship is well-documented for other arboviruses, such as DENV, where higher viremia correlates with disease severity^49–51^, yet remains largely unexplored for OROV. Notably, recent fatal OROV cases in non-endemic Brazilian regions, characterized by low Ct values (8 and 16) and rapid progression to severe hemorrhagic symptoms within days, underscore the potential link between viral load and disease severity^23^.

Phylogenetic analysis of OROV genomic sequences from Espírito Santo revealed a complex pattern of viral spread, with multiple independent introductions of the virus into the state. The samples clustered within the Brazil_2015-2024_ clade, specifically within the M_1_L_2_S_2_ reassortant lineage that caused the recent outbreak in the Amazon region^19^. The absence of monophyly in the S and M segment trees suggests that multiple introductions events occurred followed by local spread through different routes, in alignment with a recent work describing the spread of the OROV M_1_L_2_S_2_ reassortant in Brazil^22^. This molecular pattern corroborates the epidemiological data showing the initial emergence of imported cases followed by the rapid establishment of autochthonous transmission in different municipalities. Further genomic surveillance is needed to fully understand the dispersal patterns and the state’s role in the virus’s spread to other regions, highlighting the need for comprehensive and continuous surveillance throughout the region.

The predominance of OROV transmission in regions with tropical climates (Aw and Am) highlights the critical role of environmental conditions that favor a high prevalence of potential vectors, particularly *C. paraensis*. While comprehensive studies on *Culicoides* spp. distribution across Brazilian climate zones needs to be improved. European studies have documented strong associations between Köppen climate classifications and *Culicoides* spp. diversity^52^. The tropical climates characterizing the Espírito Santo outbreak zones feature high temperatures and seasonal rainfall, which likely create optimal conditions for *C. paraensis* breeding. These environments provide abundant organic matter for larval development—decomposing vegetation, fruit waste, banana tree stumps, and cacao husks, common agricultural by-products in the region, that serve as ideal breeding sites for *C. paraensis*^3,4^. Indeed, studies report peak *C. paraensis* populations during the rainy season, with temperatures between 30-32°C and relative humidity of 75-85%^53^, which present favorable macro and microclimatic conditions for vector proliferation. In contrast, temperate and subtropical highland climates, characterized by lower temperatures, more evenly distributed rainfall, and distinct vegetation, may present less favorable conditions for vector establishment. Despite the widespread distribution of *Culicoides* spp. across the Americas^30^, no cases of OROV had been reported in Espírito Santo state until 2024. The introduction of OROV into Espírito Santo is estimated to have occurred multiple times between February and March of 2024^22^, and its establishment may reflect ecological shifts that have amplified vector density or increased human-vector contact, particularly among susceptible populations in Espírito Santo and other regions of Brazil. Espírito Santo’s recent biting midge infestation in 2019^54^ likely facilitated conditions for OROV emergence by increasing vector populations in previously unaffected areas. Climate change projections in Europe suggest a shift toward warmer and wetter conditions that may transform humid subtropical climates into subtropical climates with hot summers, further promoting *Culicoides* spp. establishment^52^. Modeling studies indicate that climate-driven increases in temperature and extreme weather events could expand arboviral transmission in tropical regions like Espírito Santo, as observed for dengue^55^. These ecological changes likely interact with agricultural and land-use patterns, shaping OROV transmission and creating corridors that intensify virus-vector-host interactions. Further research is warranted to quantify the relationships between climatic variables, *C. paraensis* dynamics, and OROV transmission in Espírito Santo and other different biomes.

The significant correlations between OROV incidence and specific crops in Espírito Santo, particularly robusta coffee, cacao, coconut, and pepper plantations, underscore the complex interactions between agricultural landscapes and arboviral transmission dynamics. Outside the Amazon region, OROV transmission has been predominantly associated with rural settings, where *C. paraensis* finds optimal breeding conditions in agricultural environments. Previous studies have documented that *C. paraensis* larvae develop effectively in microhabitats created by decaying organic matter from banana and cacao plantations^3,4^, and recent outbreaks outside the Amazon have notably occurred along the Atlantic Forest biome where these crops are prevalent^22^. The emergence of OROV in Espírito Santo’s regions with high densities of robusta coffee, pepper, and coconut cultivation represents a novel association that warrants further investigation, as it broadens our understanding of potential agricultural landscapes that may support *C. paraensis* populations. This pattern aligns with the vector’s documented ability to occupy both sylvatic and anthropic environments^30^ and suggests that agricultural expansion may create ecological corridors facilitating viral spread. The fragmentation of forest landscapes by agricultural activities likely intensifies human-vector contact, particularly in areas where multiple crops create diverse microhabitats suitable for vector breeding. The strong presence of OROV in Espírito Santo’s major agricultural regions, especially where coffee and cacao cultivation predominate, emphasizes the need for detailed ecological studies of *C. paraensis* in these novel settings to identify and predict potential outbreak hotspots.

While this study offers valuable insights into the environmental and ecological factors influencing OROV emergence and spread in Espírito Santo, several limitations should be acknowledged. Inferences about spatial dissemination relied on assumptions concerning the primary vector, *C. paraensis*. However, this study did not directly assess vector competence, population density, or dispersal potential within the affected areas, which may impact the accuracy of estimates regarding transmission dynamics. Moreover, our reliance on secondary epidemiological data, rather than data from targeted field sampling, restricts the granularity and completeness of information available on transmission patterns and hotspots. The observed correlations between OROV cases and crops may also be confounded by geographical clustering effects, which were not controlled for, potentially introducing spatial biases. Future research incorporating fine-scale spatial modeling and systematic vector surveillance is essential to more precisely delineate the interactions among environmental variables, vector ecology, and OROV transmission dynamics within this emerging outbreak context.

In conclusion, our study reveals a significant epidemiological shift marked by the emergence and establishment of OROV in Espírito Santo, underscoring the virus’s capacity to adapt to new ecological landscapes outside its Amazonian origins. The rapid rise in OROV cases to levels comparable with established arboviruses like DENV and CHIKV, without apparent competitive inhibition, highlights the distinct ecological niches exploited by *Culicoides* spp. vectors in periurban and rural areas. The interplay between tropical climate, expanding agricultural landscapes, and favorable breeding sites for *C. paraensis* likely facilitated this outbreak, positioning Espírito Santo as a potential hotspot for arboviral transmission amid environmental and climatic changes. As such, our findings call for heightened surveillance of both human cases and vector populations, alongside further investigation into the eco-epidemiological drivers of OROV in Atlantic Forest regions. Understanding these dynamics will be crucial to predict, prevent, and respond to future outbreaks of OROV and other emerging arboviruses in similar environments.

## Data Availability

All data viral sequences produced in the present study, will be available in the GenBank database after the final publication.

## Funding

This study was partially funded by Fundação de Amparo à Pesquisa e Inovação do Espírito Santo-FAPES (ED and AR)

## Supplementary material

**Supplementary Methods.** Estimation of Generation Time and Calculation of the Time-Varying Reproduction Number

**Table S1.** This study utilized Oropouche samples and metadata, including their NCBI, IAL, and LACEN/ES accession numbers.

**Figure S1.** The boxplots compare the Ct values of individuals with and without each symptom. Horizontal bars represent the Ct medians and interquartile ranges (IQR). Two-sided P-values for the nonparametric Mann–Whitney test are shown for each group

## Notes

### Competing Interest Statement

The authors have declared no competing interest.

### Author Declarations

This study was approved by the Ethics Committee of the University of Vila Velha, Espírito Santo - Brazil (Certificate of Ethical Appreciation Presentation: 84698324.7.0000.5064). The Committee waived the requirement for written informed consent.

