## Supplementary material for "Oropouche Virus Outbreak in Southeast, Brazil: Expanding Beyond the Amazonian Endemic Region"

**Running title:** Oropouche Virus Outbreak in Southeast Brazil

**Author affiliations:**

[1] Laboratório de Genômica e Ecologia Viral, Centro de Ciências da Saúde, Universidade Federal do Espírito Santo, Vitória, Espírito Santo, Brazil

[2] Laboratório Central de Saúde Pública do Estado do Espírito Santo (LACEN-ES), Vitória, Espírito Santo, Brazil

[3] Instituto Oswaldo Cruz, Rio de Janeiro, Rio de Janeiro, Brazil

[4] Instituto Adolfo Lutz, São Paulo, São Paulo, Brazil

[5] Instituto Leônidas e Maria Deane, Fiocruz, Manaus, Amazonas, Brazil

[6] Laboratório de Arbovírus e Vírus Hemorrágicos, Instituto Oswaldo Cruz, Fiocruz, Rio de Janeiro, Brazil

[7] Núcleo de Doenças Infecciosas/Universidade Federal do Espírito Santo (NDI/UFES), Vitória, Espírito Santo, Brazil

### Supplementary Methods for Estimation of Generation Time and Calculation of the Time-Varying Reproduction Number

#### Natural history parameter estimates

***Human to mosquito generation time***

The human-to-mosquito generation time represents the interval between human infection and the mosquito's ability to transmit the virus through an infectious blood meal. This period encompasses the intrinsic incubation period (from infection to symptom onset) and the duration from symptom onset to viral clearance.

***Intrinsic incubation period***

While not precisely determined, the incubation period of Oropouche fever is estimated to range from 4 to 8 days in natural infections. We assumed a log-normal distribution based on Lessler et al.'s approach (1). Using the Metropolis-Hastings Markov chain Monte Carlo (MCMC) algorithm, we calibrated the model to estimate the viral incubation characteristics. We estimated a mean incubation period $\mu_{IP}$ of 6.0 (95% CrI: 5.8 – 6.3) days and a standard deviation $\sigma_{IP}$ of 1.2 (95% CrI: 1.0 – 1.5) days.

***Time to viral clearance***

The time to viral clearance refers to the duration from the onset of symptoms until the virus is no longer detectable in the blood. This period represents the window during which the virus is present in the bloodstream and may potentially be transmissible to mosquito vectors in the case of arboviruses. For OROV, observational data in humans accounts that the viral clearance period typically lasts 1-5 days, with decreasing probabilities of viral detections as the days advance (2).

Following established methods for other arboviruses (3,4), we assumed that OROV viral clearance follows a Gamma distribution with shape parameter $\alpha_C$ and scale parameter $\beta_C$, estimated using the Metropolis-Hastings MCMC algorithm. In Table A, we report the mean ($\mu_C = \alpha_C\beta_C$) and standard deviation ($\sigma_C = \alpha_C\beta_C^2$) of the time to viral clearance.

We modeled infectiousness similar to those used for Zika (3) and Mayaro (4) virus infections, with transmission beginning 1.5 days before symptom onset and ending 1.5-2 days before viral detection becomes undetectable.

By linearly scaling the time dependence by a factor $s = (\mu_{IP} - 1.5)/\mu_{IP}$, we derived the OROV human generation time, calculating its mean mean $\mu_h = s(\mu_{IP} + \mu_C)$ and standard deviation $\sigma_h = s\sqrt{\sigma_{IP}^2 + \sigma_C^2}$. to characterize the virus's transmission dynamics.

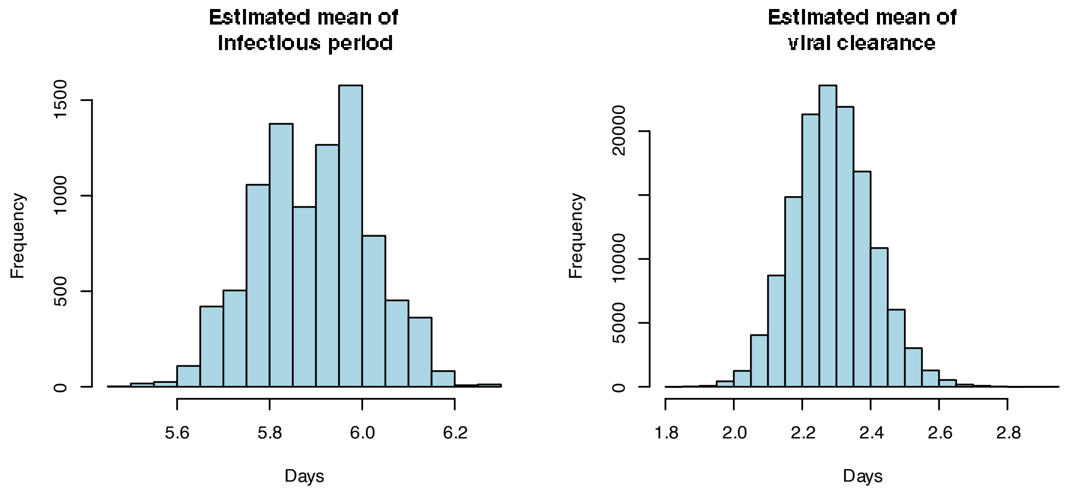


**Fig A.** Histograms of estimated means of key epidemiological parameters. The left panel represents the estimated mean of the infectious period (in days), while the right panel shows the estimated mean of viral clearance time (in days). Frequencies represent the distribution of parameter estimates obtained from the model-fitting process.

***Extrinsic incubation period***

We identified a single peer-reviewed article that investigated the susceptibility of North American mosquitoes and midges to OROV (5). This study provided sufficient information on the number of mosquitoes tested at each day post-infection, which is critical for estimating the extrinsic incubation period (EIP). The research evaluated the susceptibility of *Culex quinquefasciatus* and *Culicoides sonorensis*. Given the limited data, we aggregated information across all mosquito species and used it as a proxy for the susceptibility of *Culicoides paraensis*. The EIP was defined as the interval between infection and the dissemination of OROV to the salivary glands, rendering the mosquito capable of transmitting the virus.

We assumed that the extrinsic incubation period is Gamma distributed with shape parameter $k_{EIP}$ and scale parameter $\theta_{EIP}$, following previous studies (3,4). Using a Binomial likelihood function to estimate the probability that a mosquito is infectious by day *t*, we obtain mean posterior estimates of $k_{EIP}$ = 10.1 (95% CrI: 6.5 – 14.3) and $\theta_{EIP}$ = 1.21 (95% CrI: 0.8 – 1.7). This results in a mean EIP of 12.2 (95% CrI: 5.2 – 24.3) days with a standard deviation of 3.8 (95% CrI: 2.0 – 6.4) days (main text Table 3). Fig B shows the fitted probability density function, cumulative distribution function, and observed data.

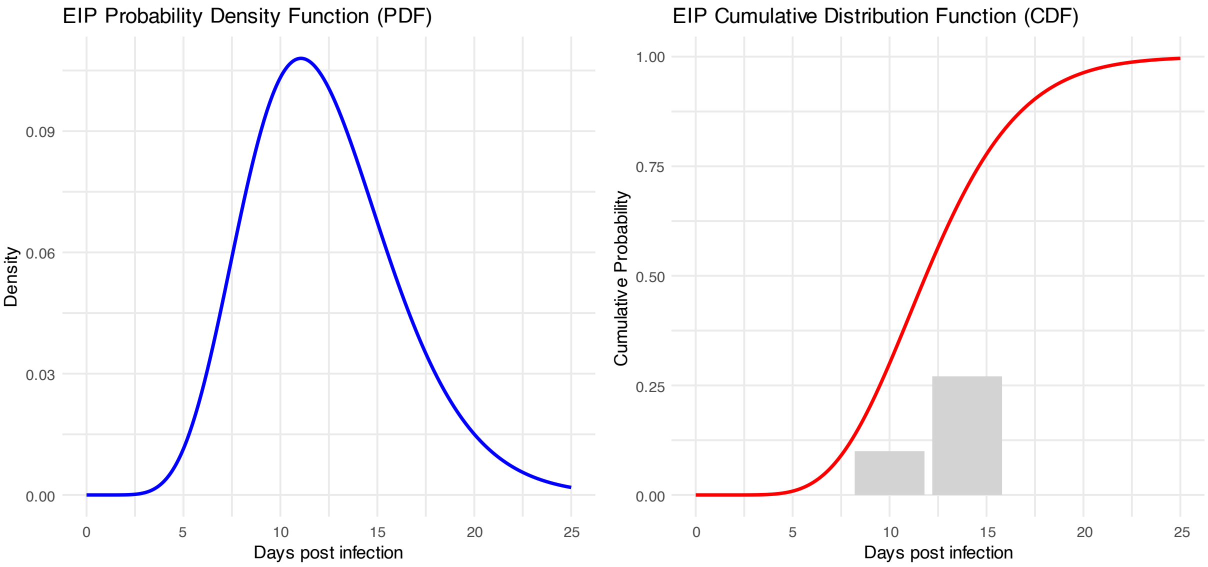


**Fig B.** Maximum likelihood EIP probability density function (left) and cumulative distribution function (right). The aggregated proportion of mosquitoes that transmitted the virus at the relative days post-infection are shown as bars.

#### *Mosquito-to-human generation time*

We followed the methodology proposed by Fergunson et al (3) and Caicedo et al. (4) to estimate the mosquito-to-human generation time, defined as the time between a vector being infected and subsequently infecting a human. However, considering the longer life cycle of *Culicoides* spp. compared to mosquitoes, we adjusted the parameters of the Gamma distribution to reflect the typical lifespan of the biting midge (6). The mosquito-to-human generation time was estimated from the estimated EIP and the mosquito daily mortality rate. We assume that the mosquito mortality rate ε is Gamma-distributed with a mean of 0.07/day and a standard deviation of 0.01/day. We estimate the mean $\mu_m$ and standard deviation $\sigma_m$ of the mosquito-to-human generation time numerically, i.e. sampling the shape parameter $k_{EIP}$ and scale parameter $\theta_{EIP}$ from their posterior distributions and ε from the Gamma distribution with mean of 0.07/day and standard deviation of 0.01/day.

We estimated a mean mosquito-to-human generation time of 11.8 (95% CrI: 5.4 – 20.9) days and a standard deviation of 4.1 (95% CrI: 3.3 – 4.9) days (Table A).

#### Generation time of OROV

Combining the estimates of the human-to-mosquito generation time with those of the mosquito-to-human generation time, we estimate that the distribution of the generation time of OROV (i.e. the time between infection of a human case and infection of the secondary human cases that case causes) has a mean of 18.0 (95% CrI: 10.7 – 28.6) days and a standard deviation of 4.3 (95% CrI: 3.4 – 5.1) days (Table A).

**Table A.** Estimates of the mean and standard deviation of the generation time distribution and its components for Oropouche virus

| Estimate | Mean (95% CrI) | Standard deviation (95% CrI) | Source |
|---|---|---|---|
| Intrinsic incubation period (days) | 6.0 (5.8-6.3) | 1.2 (1.0-1.5) | Estimated with data from (2) |
| Time to viral clearance (days) | 2.3 (1.3-3.9) | 1.3 (0.6-2.9) | Estimated with data from (2) |
| Human-to-mosquito generation time (days) | 6.2 (5.3-7.7) | 1.3 (0.9 - 2.5) | Estimated |
| Extrinsic incubation period (days) | 12.2 (6.5 - 14.3) | 1.2 (0.8 - 1.7) | Estimated with data from (5) |
| Vector lifetime (days) (*Culicoides* spp.) | 15 (fixed) | 2.5 (fixed) | From (6) |
| Mosquito-to-human generation time (days) | 11.8 (5.4-20.9) | 4.1 (3.2 - 4.9) | Estimated |
| **Oropouche virus generation time (days)** | **18.0 (10.7-28.6)** | **4.3 (3.4-5.1)** | **Estimated** |

**Estimates of the reproduction number, *R***

We estimated the instantaneous reproduction number *R* for the 2024 OROV outbreak in Espírito Santo, Brazil using the daily case counts and the generation time distribution estimated in the previous section.

The instantaneous reproduction number (*R*) was estimated using the **EpiEstim** package (7) in R (version 4.4.1) (8) within 1-week sliding time windows. For each window, *R* was computed as the median of the weekly instantaneous reproduction numbers, weighted by the weekly incidence. The resulting *R* estimates were plotted at the midpoint of their respective time windows. The estimation process employed the `uncertain_si` method from the `estimate_R()` function, incorporating a prior distribution for *R* with a mean and standard deviation of 5, as well as the mean and standard deviation of the generation time described in Table 3 of the main text.

The `uncertain_si` approach accounts for uncertainty in the serial interval distribution, as outlined by Cori et al. (7). Specifically, the mean (μ) and standard deviation (σ) of the serial interval were allowed to vary within truncated normal distributions, parameterized as shown in Table C. From these distributions, *n1* pairs of μ and σ were sampled, and for each pair, *n2* samples were drawn from the posterior distribution of *R* for each time window. Conditional on the serial interval distribution obtained, this process generated a pooled sample of size *n1*×*n2*, representing the joint posterior distribution of *R* across all time windows.

**Table B. Values used in estimate_R() function in EpiEstim package.**

| Parameter | Days |
|---|---|
| **mean_si** | 18.0 |
| **std_mean_si** | 1.0 |
| **min_mean_si** | 10.7 |
| **max_mean_si** | 28.6 |
| **std_si** | 4.3 |
| **std_std_si** | 0.43 |
| **min_std_si** | 3.4 |
| **max_std_si** | 5.1 |

**Table S1.** This study utilized Oropouche samples and metadata, including their NCBI, IAL, and LACEN/ES accession numbers.

| NCBI/IAL/LACEN number access | Genome segment | Collection date (yyyy-mm-dd) | Location | Major groups classification |
|---|---|---|---|---|
| KP026179.1 | long | 1955-00-00 | Trinidad and Tobago_SangreGrande | TT_1955 |
| KP026180.1 | medium | 1955-00-00 | Trinidad and Tobago_SangreGrande | TT_1955 |
| KP026181.1 | small | 1955-00-00 | Trinidad and Tobago_SangreGrande | TT_1955 |
| KP052850.1 | long | 1960-00-00 | Brazil | OROV prototype |
| KP052851.1 | medium | 1960-00-00 | Brazil | OROV prototype |
| KP052852.1 | small | 1960-00-00 | Brazil | OROV prototype |
| MG747524.1 | small | 1960-00-00 | Brazil | BR_1960-2006 |
| MG747525.1 | medium | 1960-00-00 | Brazil | BR_1960-2006 |
| MG747526.1 | long | 1960-00-00 | Brazil | BR_1960-2006 |
| MG747527.1 | small | 1961-00-00 | Brazil | BR_1960-2006 |
| MG747528.1 | medium | 1961-00-00 | Brazil | BR_1960-2006 |
| MG747529.1 | long | 1961-00-00 | Brazil | BR_1960-2006 |
| MG747530.1 | small | 1961-00-00 | Brazil | BR_1960-2006 |
| MG747531.1 | medium | 1961-00-00 | Brazil | BR_1960-2006 |
| MG747532.1 | long | 1961-00-00 | Brazil | BR_1960-2006 |
| MG747533.1 | small | 1967-00-00 | Brazil | BR_1960-2006 |
| MG747534.1 | medium | 1967-00-00 | Brazil | BR_1960-2006 |
| MG747535.1 | long | 1967-00-00 | Brazil | BR_1960-2006 |
| MG747536.1 | small | 1968-00-00 | Brazil | BR_1960-2006 |
| MG747537.1 | medium | 1968-00-00 | Brazil | BR_1960-2006 |
| MG747538.1 | long | 1968-00-00 | Brazil | BR_1960-2006 |
| MG747539.1 | small | 1971-00-00 | Brazil | BR_1960-2006 |
| MG747540.1 | medium | 1971-00-00 | Brazil | BR_1960-2006 |
| MG747541.1 | long | 1971-00-00 | Brazil | BR_1960-2006 |
| MG747542.1 | small | 1971-00-00 | Brazil | BR_1960-2006 |
| MG747543.1 | medium | 1971-00-00 | Brazil | BR_1960-2006 |
| MG747544.1 | long | 1971-00-00 | Brazil | BR_1960-2006 |
| MG747545.1 | small | 1971-00-00 | Brazil | BR_1960-2006 |
| MG747546.1 | medium | 1971-00-00 | Brazil | BR_1960-2006 |
| MG747547.1 | long | 1971-00-00 | Brazil | BR_1960-2006 |
| MG747548.1 | small | 1978-00-00 | Brazil | BR_1960-2006 |
| MG747549.1 | medium | 1978-00-00 | Brazil | BR_1960-2006 |
| MG747550.1 | long | 1978-00-00 | Brazil | BR_1960-2006 |
| MG747551.1 | small | 1979-00-00 | Brazil | BR_1960-2006 |
| MG747552.1 | medium | 1979-00-00 | Brazil | BR_1960-2006 |
| MG747553.1 | long | 1979-00-00 | Brazil | BR_1960-2006 |
| MG747506.1 | small | 1980-00-00 | Brazil | BR_1960-2006 |

| | | | | |
|---|---|---|---|---|
| MG747507.1 | medium | 1980-00-00 | Brazil | BR_1960-2006 |
| MG747508.1 | long | 1980-00-00 | Brazil | BR_1960-2006 |
| MG747509.1 | small | 1980-00-00 | Brazil | BR_1960-2006 |
| MG747510.1 | medium | 1980-00-00 | Brazil | BR_1960-2006 |
| MG747511.1 | long | 1980-00-00 | Brazil | BR_1960-2006 |
| MG747554.1 | small | 1980-00-00 | Brazil | BR_1960-2006 |
| MG747555.1 | medium | 1980-00-00 | Brazil | BR_1960-2006 |
| MG747556.1 | long | 1980-00-00 | Brazil | BR_1960-2006 |
| MG747512.1 | small | 1988-00-00 | Brazil | BR_1960-2006 |
| MG747513.1 | medium | 1988-00-00 | Brazil | BR_1960-2006 |
| MG747514.1 | long | 1988-00-00 | Brazil | BR_1960-2006 |
| MG747515.1 | small | 1988-00-00 | Brazil | BR_1960-2006 |
| MG747516.1 | medium | 1988-00-00 | Brazil | BR_1960-2006 |
| MG747517.1 | long | 1988-00-00 | Brazil | BR_1960-2006 |
| KP795075.1 | long | 1989-00-00 | Panama_SanMiguelito | Panama_1989-1999 |
| KP795076.1 | medium | 1989-00-00 | Panama_SanMiguelito | Panama_1989-1999 |
| KP795077.1 | small | 1989-00-00 | Panama_SanMiguelito | Panama_1989-1999 |
| KP795081.1 | long | 1989-00-00 | Panama_CidadedoPanama | Panama_1989-1999 |
| KP795082.1 | medium | 1989-00-00 | Panama_CidadedoPanama | Panama_1989-1999 |
| KP795083.1 | small | 1989-00-00 | Panama_CidadedoPanama | Panama_1989-1999 |
| KP795078.1 | long | 1989-08-00 | Panama | Panama_1989-1999 |
| KP795079.1 | medium | 1989-08-00 | Panama | Panama_1989-1999 |
| KP795080.1 | small | 1989-08-00 | Panama | Panama_1989-1999 |
| MG747602.1 | small | 1990-00-00 | Brazil | BR_1960-2006 |
| MG747603.1 | medium | 1990-00-00 | Brazil | BR_1960-2006 |
| MG747604.1 | long | 1990-00-00 | Brazil | BR_1960-2006 |
| MG747605.1 | small | 1991-00-00 | Brazil | BR_1960-2006 |
| MG747606.1 | medium | 1991-00-00 | Brazil | BR_1960-2006 |
| MG747607.1 | long | 1991-00-00 | Brazil | BR_1960-2006 |
| PP357048.1 | long | 1991-00-00 | Brazil_Rondonia | BR_1960-2006 |
| PP357049.1 | medium | 1991-00-00 | Brazil_Rondonia | BR_1960-2006 |
| PP357050.1 | small | 1991-00-00 | Brazil_Rondonia | BR_1960-2006 |
| KP795072.1 | long | 1992-04-00 | Peru | Peru_1992-2000 |
| KP795073.1 | medium | 1992-04-00 | Peru | Peru_1992-2000 |
| KP795074.1 | small | 1992-04-00 | Peru | Peru_1992-2000 |
| MG747518.1 | small | 1993-00-00 | Brazil | BR_1960-2006 |
| MG747519.1 | medium | 1993-00-00 | Brazil | BR_1960-2006 |
| MG747520.1 | long | 1993-00-00 | Brazil | BR_1960-2006 |
| MG747557.1 | small | 1994-00-00 | Brazil | BR_1960-2006 |

| | | | | |
|---|---|---|---|---|
| MG747558.1 | medium | 1994-00-00 | Brazil | BR_1960-2006 |
| MG747559.1 | long | 1994-00-00 | Brazil | BR_1960-2006 |
| MG747560.1 | small | 1994-00-00 | Brazil | BR_1960-2006 |
| MG747561.1 | medium | 1994-00-00 | Brazil | BR_1960-2006 |
| MG747562.1 | long | 1994-00-00 | Brazil | BR_1960-2006 |
| MG747563.1 | small | 1994-00-00 | Brazil | BR_1960-2006 |
| MG747564.1 | medium | 1994-00-00 | Brazil | BR_1960-2006 |
| MG747565.1 | long | 1994-00-00 | Brazil | BR_1960-2006 |
| MG747566.1 | small | 1994-00-00 | Brazil | BR_1960-2006 |
| MG747567.1 | medium | 1994-00-00 | Brazil | BR_1960-2006 |
| MG747568.1 | long | 1994-00-00 | Brazil | BR_1960-2006 |
| MG747569.1 | small | 1994-00-00 | Brazil | BR_1960-2006 |
| MG747570.1 | medium | 1994-00-00 | Brazil | BR_1960-2006 |
| MG747571.1 | long | 1994-00-00 | Brazil | BR_1960-2006 |
| KP795096.1 | long | 1994-02-00 | Peru | Peru_1992-2000 |
| KP795097.1 | medium | 1994-02-00 | Peru | Peru_1992-2000 |
| KP795098.1 | small | 1994-02-00 | Peru | Peru_1992-2000 |
| KP795087.1 | long | 1995-06-00 | Peru | Peru_1992-2000 |
| KP795088.1 | medium | 1995-06-00 | Peru | Peru_1992-2000 |
| KP795089.1 | small | 1995-06-00 | Peru | Peru_1992-2000 |
| MG747503.1 | small | 1996-00-00 | Brazil | BR_1960-2006 |
| MG747504.1 | medium | 1996-00-00 | Brazil | BR_1960-2006 |
| MG747505.1 | long | 1996-00-00 | Brazil | BR_1960-2006 |
| MG747572.1 | small | 1996-00-00 | Brazil | BR_1960-2006 |
| MG747573.1 | medium | 1996-00-00 | Brazil | BR_1960-2006 |
| MG747574.1 | long | 1996-00-00 | Brazil | BR_1960-2006 |
| MG747575.1 | small | 1996-00-00 | Brazil | BR_1960-2006 |
| MG747576.1 | medium | 1996-00-00 | Brazil | BR_1960-2006 |
| MG747577.1 | long | 1996-00-00 | Brazil | BR_1960-2006 |
| MG747578.1 | small | 1996-00-00 | Brazil | BR_1960-2006 |
| MG747579.1 | medium | 1996-00-00 | Brazil | BR_1960-2006 |
| MG747580.1 | long | 1996-00-00 | Brazil | BR_1960-2006 |
| KP795090.1 | long | 1997-08-00 | Peru | Peru_1992-2000 |
| KP795091.1 | medium | 1997-08-00 | Peru | Peru_1992-2000 |
| KP795092.1 | small | 1997-08-00 | Peru | Peru_1992-2000 |
| KP795093.1 | long | 1998-04-00 | Peru | Peru_1992-2000 |
| KP795094.1 | medium | 1998-04-00 | Peru | Peru_1992-2000 |
| KP795095.1 | small | 1998-04-00 | Peru | Peru_1992-2000 |
| KF697142.1 | long | 1999-00-00 | Peru_Iquitos | IQTV prototype |
| KF697143.1 | medium | 1999-00-00 | Peru_Iquitos | IQTV prototype |
| KF697144.1 | small | 1999-00-00 | Peru_Iquitos | IQTV prototype |

| | | | | |
|---|---|---|---|---|
| KP795102.1 | long | 1999-10-00 | Panama | Panama_1989-1999 |
| KP795103.1 | medium | 1999-10-00 | Panama | Panama_1989-1999 |
| KP795104.1 | small | 1999-10-00 | Panama | Panama_1989-1999 |
| MG747521.1 | small | 2000-00-00 | Brazil | BR_1960-2006 |
| MG747522.1 | medium | 2000-00-00 | Brazil | BR_1960-2006 |
| MG747523.1 | long | 2000-00-00 | Brazil | BR_1960-2006 |
| KP795099.1 | long | 2000-05-00 | Peru | Peru_1992-2000 |
| KP795100.1 | medium | 2000-05-00 | Peru | Peru_1992-2000 |
| KP795101.1 | small | 2000-05-00 | Peru | Peru_1992-2000 |
| MG747581.1 | small | 2003-00-00 | Brazil | BR_1960-2006 |
| MG747582.1 | medium | 2003-00-00 | Brazil | BR_1960-2006 |
| MG747583.1 | long | 2003-00-00 | Brazil | BR_1960-2006 |
| MG747584.1 | small | 2003-00-00 | Brazil | BR_1960-2006 |
| MG747585.1 | medium | 2003-00-00 | Brazil | BR_1960-2006 |
| MG747586.1 | long | 2003-00-00 | Brazil | BR_1960-2006 |
| MG747587.1 | small | 2004-00-00 | Brazil | BR_1960-2006 |
| MG747588.1 | medium | 2004-00-00 | Brazil | BR_1960-2006 |
| MG747589.1 | long | 2004-00-00 | Brazil | BR_1960-2006 |
| MG747590.1 | small | 2004-00-00 | Brazil | BR_1960-2006 |
| MG747591.1 | medium | 2004-00-00 | Brazil | BR_1960-2006 |
| MG747592.1 | long | 2004-00-00 | Brazil | BR_1960-2006 |
| MG747593.1 | small | 2006-00-00 | Brazil | BR_1960-2006 |
| MG747594.1 | medium | 2006-00-00 | Brazil | BR_1960-2006 |
| MG747595.1 | long | 2006-00-00 | Brazil | BR_1960-2006 |
| MG747596.1 | small | 2006-00-00 | Brazil | BR_1960-2006 |
| MG747597.1 | medium | 2006-00-00 | Brazil | BR_1960-2006 |
| MG747598.1 | long | 2006-00-00 | Brazil | BR_1960-2006 |
| MG747599.1 | small | 2006-00-00 | Brazil | BR_1960-2006 |
| MG747600.1 | medium | 2006-00-00 | Brazil | BR_1960-2006 |
| MG747601.1 | long | 2006-00-00 | Brazil | BR_1960-2006 |
| KF697145.1 | medium | 2007-00-00 | Peru_MadreDios | MDDV prototype |
| KF697146.1 | small | 2007-00-00 | Peru_MadreDios | MDDV prototype |
| KF697147.1 | long | 2007-00-00 | Peru_MadreDios | MDDV prototype |
| KP795084.1 | long | 2008-03-00 | Peru | Peru-Ecuador_2008-2016 |
| KP795085.1 | medium | 2008-03-00 | Peru | Peru-Ecuador_2008-2016 |
| KP795086.1 | small | 2008-03-00 | Peru | Peru-Ecuador_2008-2016 |
| KP691618.1 | long | 2009-06-00 | Brazil | BR_2009-2018 |
| KP691619.1 | medium | 2009-06-00 | Brazil | BR_2009-2018 |

| | | | | |
|---|---|---|---|---|
| KP691620.1 | small | 2009-06-00 | Brazil | BR_2009-2018 |
| KP691621.1 | long | 2009-06-00 | Brazil | BR_2009-2018 |
| KP691622.1 | medium | 2009-06-00 | Brazil | BR_2009-2018 |
| KP691623.1 | small | 2009-06-00 | Brazil | BR_2009-2018 |
| OP407852.1 | long | 2009-06-00 | Brazil | BR_2009-2018 |
| OP407853.1 | medium | 2009-06-00 | Brazil | BR_2009-2018 |
| OP407854.1 | small | 2009-06-00 | Brazil | BR_2009-2018 |
| KP691603.1 | long | 2009-07-00 | Brazil | BR_2009-2018 |
| KP691604.1 | medium | 2009-07-00 | Brazil | BR_2009-2018 |
| KP691605.1 | small | 2009-07-00 | Brazil | BR_2009-2018 |
| KP691606.1 | long | 2009-07-00 | Brazil | BR_2009-2018 |
| KP691607.1 | medium | 2009-07-00 | Brazil | BR_2009-2018 |
| KP691608.1 | small | 2009-07-00 | Brazil | BR_2009-2018 |
| KP691609.1 | long | 2009-07-00 | Brazil | BR_2009-2018 |
| KP691610.1 | medium | 2009-07-00 | Brazil | BR_2009-2018 |
| KP691611.1 | small | 2009-07-00 | Brazil | BR_2009-2018 |
| KP691612.1 | long | 2009-07-00 | Brazil | BR_2009-2018 |
| KP691613.1 | medium | 2009-07-00 | Brazil | BR_2009-2018 |
| KP691614.1 | small | 2009-07-00 | Brazil | BR_2009-2018 |
| KP691615.1 | long | 2009-07-00 | Brazil | BR_2009-2018 |
| KP691616.1 | medium | 2009-07-00 | Brazil | BR_2009-2018 |
| KP691617.1 | small | 2009-07-00 | Brazil | BR_2009-2018 |
| KP691630.1 | long | 2009-08-00 | Brazil | BR_2009-2018 |
| KP691631.1 | medium | 2009-08-00 | Brazil | BR_2009-2018 |
| KP691632.1 | small | 2009-08-00 | Brazil | BR_2009-2018 |
| KP691627.1 | long | 2012-00-00 | Brazil_MinasGerais_Perdoes | PEDV prototype |
| KP691628.1 | medium | 2012-00-00 | Brazil_MinasGerais_Perdoes | PEDV prototype |
| KP691629.1 | small | 2012-00-00 | Brazil_MinasGerais_Perdoes | PEDV prototype |
| MN264267.1 | long | 2014-05-00 | Haiti | Haiti_2014 |
| MN264268.1 | medium | 2014-05-00 | Haiti | Haiti_2014 |
| MN264269.1 | small | 2014-05-00 | Haiti | Haiti_2014 |
| PP154170.1 | small | 2015-04-00 | Brazil_Amazonas_Tefe | BR_2015-2023 |
| PP154171.1 | medium | 2015-04-00 | Brazil_Amazonas_Tefe | BR_2015-2023 |
| PP154172.1 | long | 2015-04-00 | Brazil_Amazonas_Tefe | BR_2015-2023 |
| MF926352.1 | small | 2016-04-00 | Ecuador | Peru-Ecuador_2008-2016 |
| MF926353.1 | medium | 2016-04-00 | Ecuador | Peru-Ecuador_2008-2016 |
| MF926354.1 | long | 2016-04-00 | Ecuador | Peru-Ecuador_2008-2016 |
| MK506818.1 | small | 2016-04-00 | Ecuador | Peru-Ecuador_2008-2016 |

| | | | | |
|---|---|---|---|---|
| MK506819.1 | small | 2016-04-00 | Ecuador | Peru-Ecuador_2008-2016 |
| MK506820.1 | small | 2016-04-00 | Ecuador | Peru-Ecuador_2008-2016 |
| MK506821.1 | small | 2016-04-00 | Ecuador | Peru-Ecuador_2008-2016 |
| MK506822.1 | small | 2016-04-00 | Ecuador | Peru-Ecuador_2008-2016 |
| MK506823.1 | medium | 2016-04-00 | Ecuador | Peru-Ecuador_2008-2016 |
| MK506824.1 | medium | 2016-04-00 | Ecuador | Peru-Ecuador_2008-2016 |
| MK506825.1 | medium | 2016-04-00 | Ecuador | Peru-Ecuador_2008-2016 |
| MK506826.1 | medium | 2016-04-00 | Ecuador | Peru-Ecuador_2008-2016 |
| MK506827.1 | medium | 2016-04-00 | Ecuador | Peru-Ecuador_2008-2016 |
| MK506828.1 | long | 2016-04-00 | Ecuador | Peru-Ecuador_2008-2016 |
| MK506829.1 | long | 2016-04-00 | Ecuador | Peru-Ecuador_2008-2016 |
| MK506830.1 | long | 2016-04-00 | Ecuador | Peru-Ecuador_2008-2016 |
| MK506831.1 | long | 2016-04-00 | Ecuador | Peru-Ecuador_2008-2016 |
| MK506832.1 | long | 2016-04-00 | Ecuador | Peru-Ecuador_2008-2016 |
| MT879228.1 | long | 2018-03-00 | Brazil | BR_2009-2018 |
| MT879229.1 | medium | 2018-03-00 | Brazil | BR_2009-2018 |
| MT879230.1 | small | 2018-03-00 | Brazil | BR_2009-2018 |
| PP153981.1 | small | 2022-08-00 | Brazil_Roraima_AltoAlegre | BR_2015-2023 |
| PP153982.1 | medium | 2022-08-00 | Brazil_Roraima_AltoAlegre | BR_2015-2023 |
| PP153983.1 | long | 2022-08-00 | Brazil_Roraima_AltoAlegre | BR_2015-2023 |
| PP153977.1 | small | 2022-10-00 | Brazil_Rondonia_PortoVelho | BR_2015-2023 |
| PP153978.1 | medium | 2022-10-00 | Brazil_Rondonia_PortoVelho | BR_2015-2023 |
| PP153979.1 | long | 2022-10-00 | Brazil_Rondonia_PortoVelho | BR_2015-2023 |
| PP154128.1 | small | 2022-12-00 | Brazil_Amazonas_Manaus | BR_2015-2023 |
| PP154129.1 | medium | 2022-12-00 | Brazil_Amazonas_Manaus | BR_2015-2023 |
| PP154130.1 | long | 2022-12-00 | Brazil_Amazonas_Manaus | BR_2015-2023 |
| PP153951.1 | small | 2023-01-00 | Brazil_Rondonia_PortoVelho | BR_2015-2023 |
| PP153952.1 | medium | 2023-01-00 | Brazil_Rondonia_PortoVelho | BR_2015-2023 |
| PP153953.1 | long | 2023-01-00 | Brazil_Rondonia_PortoVelho | BR_2015-2023 |

| | | | | |
|---|---|---|---|---|
| PP153975.1 | small | 2023-01-00 | Brazil_Rondonia_PortoVelho | BR_2015-2023 |
| PP153976.1 | long | 2023-01-00 | Brazil_Rondonia_PortoVelho | BR_2015-2023 |
| PP153980.1 | medium | 2023-01-00 | Brazil_Rondonia_PortoVelho | BR_2015-2023 |
| PP154011.1 | small | 2023-01-00 | Brazil_Roraima_SaoJoaoBaliza | BR_2015-2023 |
| PP154012.1 | medium | 2023-01-00 | Brazil_Roraima_SaoJoaoBaliza | BR_2015-2023 |
| PP154013.1 | long | 2023-01-00 | Brazil_Roraima_SaoJoaoBaliza | BR_2015-2023 |
| PP154146.1 | small | 2023-01-00 | Brazil_Amazonas_Labrea | BR_2015-2023 |
| PP154147.1 | medium | 2023-01-00 | Brazil_Amazonas_Labrea | BR_2015-2023 |
| PP154148.1 | long | 2023-01-00 | Brazil_Amazonas_Labrea | BR_2015-2023 |
| PP154149.1 | small | 2023-01-00 | Brazil_Amazonas_Manicore | BR_2015-2023 |
| PP154150.1 | medium | 2023-01-00 | Brazil_Amazonas_Manicore | BR_2015-2023 |
| PP154151.1 | long | 2023-01-00 | Brazil_Amazonas_Manicore | BR_2015-2023 |
| PP153966.1 | small | 2023-02-00 | Brazil_Rondonia_PortoVelho | BR_2015-2023 |
| PP153967.1 | medium | 2023-02-00 | Brazil_Rondonia_PortoVelho | BR_2015-2023 |
| PP153968.1 | long | 2023-02-00 | Brazil_Rondonia_PortoVelho | BR_2015-2023 |
| PP153969.1 | small | 2023-02-00 | Brazil_Rondonia_PortoVelho | BR_2015-2023 |
| PP153970.1 | medium | 2023-02-00 | Brazil_Rondonia_PortoVelho | BR_2015-2023 |
| PP153971.1 | long | 2023-02-00 | Brazil_Rondonia_PortoVelho | BR_2015-2023 |
| PP153972.1 | small | 2023-02-00 | Brazil_Rondonia_PortoVelho | BR_2015-2023 |
| PP153973.1 | medium | 2023-02-00 | Brazil_Rondonia_PortoVelho | BR_2015-2023 |
| PP153974.1 | long | 2023-02-00 | Brazil_Rondonia_PortoVelho | BR_2015-2023 |
| PP153993.1 | small | 2023-02-00 | Brazil_Roraima_Rorainopolis | BR_2015-2023 |
| PP153994.1 | medium | 2023-02-00 | Brazil_Roraima_Rorainopolis | BR_2015-2023 |
| PP153995.1 | long | 2023-02-00 | Brazil_Roraima_Rorainopolis | BR_2015-2023 |
| PP153996.1 | medium | 2023-02-00 | Brazil_Roraima_SaoJoaoBaliza | BR_2015-2023 |
| PP153997.1 | small | 2023-02-00 | Brazil_Roraima_SaoJoaoBaliza | BR_2015-2023 |
| PP153998.1 | long | 2023-02-00 | Brazil_Roraima_SaoJoaoBaliza | BR_2015-2023 |
| PP153999.1 | small | 2023-02-00 | Brazil_Roraima_Rorainopolis | BR_2015-2023 |
| PP154000.1 | medium | 2023-02-00 | Brazil_Roraima_Rorainopolis | BR_2015-2023 |
| PP154001.1 | long | 2023-02-00 | Brazil_Roraima_Rorainopolis | BR_2015-2023 |
| PP154002.1 | small | 2023-02-00 | Brazil_Roraima_Rorainopolis | BR_2015-2023 |
| PP154003.1 | medium | 2023-02-00 | Brazil_Roraima_Rorainopolis | BR_2015-2023 |
| PP154004.1 | long | 2023-02-00 | Brazil_Roraima_Rorainopolis | BR_2015-2023 |
| PP154005.1 | small | 2023-02-00 | Brazil_Roraima_Rorainopolis | BR_2015-2023 |
| PP154006.1 | medium | 2023-02-00 | Brazil_Roraima_Rorainopolis | BR_2015-2023 |
| PP154007.1 | long | 2023-02-00 | Brazil_Roraima_Rorainopolis | BR_2015-2023 |
| PP154008.1 | small | 2023-02-00 | Brazil_Roraima_Rorainopolis | BR_2015-2023 |
| PP154009.1 | medium | 2023-02-00 | Brazil_Roraima_Rorainopolis | BR_2015-2023 |
| PP154010.1 | long | 2023-02-00 | Brazil_Roraima_Rorainopolis | BR_2015-2023 |
| PP154014.1 | small | 2023-02-00 | Brazil_Roraima_Canta | BR_2015-2023 |
| PP154015.1 | medium | 2023-02-00 | Brazil_Roraima_Canta | BR_2015-2023 |
| PP154016.1 | long | 2023-02-00 | Brazil_Roraima_Canta | BR_2015-2023 |

| | | | | |
|---|---|---|---|---|
| PP154131.1 | small | 2023-02-00 | Brazil_Amazonas_Labrea | BR_2015-2023 |
| PP154132.1 | medium | 2023-02-00 | Brazil_Amazonas_Labrea | BR_2015-2023 |
| PP154133.1 | long | 2023-02-00 | Brazil_Amazonas_Labrea | BR_2015-2023 |
| PP154134.1 | small | 2023-02-00 | Brazil_Amazonas_Labrea | BR_2015-2023 |
| PP154135.1 | medium | 2023-02-00 | Brazil_Amazonas_Labrea | BR_2015-2023 |
| PP154136.1 | long | 2023-02-00 | Brazil_Amazonas_Labrea | BR_2015-2023 |
| PP154137.1 | small | 2023-02-00 | Brazil_Amazonas_Labrea | BR_2015-2023 |
| PP154138.1 | medium | 2023-02-00 | Brazil_Amazonas_Labrea | BR_2015-2023 |
| PP154139.1 | long | 2023-02-00 | Brazil_Amazonas_Labrea | BR_2015-2023 |
| PP154140.1 | small | 2023-02-00 | Brazil_Amazonas_Manicore | BR_2015-2023 |
| PP154141.1 | medium | 2023-02-00 | Brazil_Amazonas_Manicore | BR_2015-2023 |
| PP154142.1 | long | 2023-02-00 | Brazil_Amazonas_Manicore | BR_2015-2023 |
| PP154143.1 | small | 2023-02-00 | Brazil_Amazonas_Manicore | BR_2015-2023 |
| PP154144.1 | medium | 2023-02-00 | Brazil_Amazonas_Manicore | BR_2015-2023 |
| PP154145.1 | long | 2023-02-00 | Brazil_Amazonas_Manicore | BR_2015-2023 |
| PP153948.1 | small | 2023-03-00 | Brazil_Rondonia_PortoVelho | BR_2015-2023 |
| PP153949.1 | medium | 2023-03-00 | Brazil_Rondonia_PortoVelho | BR_2015-2023 |
| PP153950.1 | long | 2023-03-00 | Brazil_Rondonia_PortoVelho | BR_2015-2023 |
| PP153954.1 | small | 2023-03-00 | Brazil_Rondonia_PortoVelho | BR_2015-2023 |
| PP153955.1 | medium | 2023-03-00 | Brazil_Rondonia_PortoVelho | BR_2015-2023 |
| PP153956.1 | long | 2023-03-00 | Brazil_Rondonia_PortoVelho | BR_2015-2023 |
| PP153957.1 | small | 2023-03-00 | Brazil_Rondonia_PortoVelho | BR_2015-2023 |
| PP153958.1 | medium | 2023-03-00 | Brazil_Rondonia_PortoVelho | BR_2015-2023 |
| PP153959.1 | long | 2023-03-00 | Brazil_Rondonia_PortoVelho | BR_2015-2023 |
| PP153960.1 | small | 2023-03-00 | Brazil_Rondonia_PortoVelho | BR_2015-2023 |
| PP153961.1 | medium | 2023-03-00 | Brazil_Rondonia_PortoVelho | BR_2015-2023 |
| PP153962.1 | long | 2023-03-00 | Brazil_Rondonia_PortoVelho | BR_2015-2023 |
| PP153963.1 | small | 2023-03-00 | Brazil_Rondonia_PortoVelho | BR_2015-2023 |
| PP153964.1 | medium | 2023-03-00 | Brazil_Rondonia_PortoVelho | BR_2015-2023 |
| PP153965.1 | long | 2023-03-00 | Brazil_Rondonia_PortoVelho | BR_2015-2023 |
| PP154017.1 | small | 2023-03-00 | Brazil_Rondonia_ColoradodoOeste | BR_2015-2023 |
| PP154018.1 | medium | 2023-03-00 | Brazil_Rondonia_ColoradodoOeste | BR_2015-2023 |
| PP154019.1 | long | 2023-03-00 | Brazil_Rondonia_ColoradodoOeste | BR_2015-2023 |
| PP154023.1 | small | 2023-03-00 | Brazil_Rondonia_MonteNegro | BR_2015-2023 |
| PP154024.1 | medium | 2023-03-00 | Brazil_Rondonia_MonteNegro | BR_2015-2023 |
| PP154025.1 | long | 2023-03-00 | Brazil_Rondonia_MonteNegro | BR_2015-2023 |
| PP154026.1 | small | 2023-03-00 | Brazil_Rondonia_MonteNegro | BR_2015-2023 |
| PP154027.1 | medium | 2023-03-00 | Brazil_Rondonia_MonteNegro | BR_2015-2023 |
| PP154028.1 | long | 2023-03-00 | Brazil_Rondonia_MonteNegro | BR_2015-2023 |
| PP154029.1 | small | 2023-03-00 | Brazil_Rondonia_Corumbiara | BR_2015-2023 |
| PP154030.1 | medium | 2023-03-00 | Brazil_Rondonia_Corumbiara | BR_2015-2023 |
| PP154031.1 | long | 2023-03-00 | Brazil_Rondonia_Corumbiara | BR_2015-2023 |

| | | | | |
|---|---|---|---|---|
| PP154032.1 | small | 2023-03-00 | Brazil_Rondonia_Corumbiara | BR_2015-2023 |
| PP154033.1 | medium | 2023-03-00 | Brazil_Rondonia_Corumbiara | BR_2015-2023 |
| PP154034.1 | long | 2023-03-00 | Brazil_Rondonia_Corumbiara | BR_2015-2023 |
| PP154152.1 | small | 2023-03-00 | Brazil_Acre_RioBranco | BR_2015-2023 |
| PP154153.1 | medium | 2023-03-00 | Brazil_Acre_RioBranco | BR_2015-2023 |
| PP154154.1 | long | 2023-03-00 | Brazil_Acre_RioBranco | BR_2015-2023 |
| PP154155.1 | small | 2023-03-00 | Brazil_Acre_RioBranco | BR_2015-2023 |
| PP154156.1 | medium | 2023-03-00 | Brazil_Acre_RioBranco | BR_2015-2023 |
| PP154157.1 | long | 2023-03-00 | Brazil_Acre_RioBranco | BR_2015-2023 |
| PP154158.1 | small | 2023-03-00 | Brazil_Acre_PortoAcre | BR_2015-2023 |
| PP154159.1 | medium | 2023-03-00 | Brazil_Acre_PortoAcre | BR_2015-2023 |
| PP154160.1 | long | 2023-03-00 | Brazil_Acre_PortoAcre | BR_2015-2023 |
| PP154161.1 | small | 2023-03-00 | Brazil_Acre_Acrelandia | BR_2015-2023 |
| PP154162.1 | medium | 2023-03-00 | Brazil_Acre_Acrelandia | BR_2015-2023 |
| PP154163.1 | long | 2023-03-00 | Brazil_Acre_Acrelandia | BR_2015-2023 |
| PP154164.1 | small | 2023-03-00 | Brazil_Acre_Acrelandia | BR_2015-2023 |
| PP154165.1 | medium | 2023-03-00 | Brazil_Acre_Acrelandia | BR_2015-2023 |
| PP154166.1 | long | 2023-03-00 | Brazil_Acre_Acrelandia | BR_2015-2023 |
| PP154167.1 | small | 2023-03-00 | Brazil_Acre_Acrelandia | BR_2015-2023 |
| PP154168.1 | medium | 2023-03-00 | Brazil_Acre_Acrelandia | BR_2015-2023 |
| PP154169.1 | long | 2023-03-00 | Brazil_Acre_Acrelandia | BR_2015-2023 |
| PP154020.1 | small | 2023-04-00 | Brazil_Rondonia_CostaMarques | BR_2015-2023 |
| PP154021.1 | medium | 2023-04-00 | Brazil_Rondonia_CostaMarques | BR_2015-2023 |
| PP154022.1 | long | 2023-04-00 | Brazil_Rondonia_CostaMarques | BR_2015-2023 |
| PP154035.1 | small | 2023-04-00 | Brazil_Rondonia_SaoFranciscodoGuapore | BR_2015-2023 |
| PP154036.1 | medium | 2023-04-00 | Brazil_Rondonia_SaoFranciscodoGuapore | BR_2015-2023 |
| PP154037.1 | long | 2023-04-00 | Brazil_Rondonia_SaoFranciscodoGuapore | BR_2015-2023 |
| PP153945.1 | small | 2023-05-00 | Brazil_Rondonia_PortoVelho | BR_2015-2023 |
| PP153946.1 | medium | 2023-05-00 | Brazil_Rondonia_PortoVelho | BR_2015-2023 |
| PP153947.1 | long | 2023-05-00 | Brazil_Rondonia_PortoVelho | BR_2015-2023 |
| PP154038.1 | small | 2023-05-00 | Brazil_Rondonia_Cacaulandia | BR_2015-2023 |
| PP154039.1 | medium | 2023-05-00 | Brazil_Rondonia_Cacaulandia | BR_2015-2023 |
| PP154040.1 | long | 2023-05-00 | Brazil_Rondonia_Cacaulandia | BR_2015-2023 |
| PP153984.1 | small | 2023-08-00 | Brazil_Roraima_Mucajai | BR_2015-2023 |
| PP153985.1 | medium | 2023-08-00 | Brazil_Roraima_Mucajai | BR_2015-2023 |
| PP153986.1 | long | 2023-08-00 | Brazil_Roraima_Mucajai | BR_2015-2023 |
| PP153987.1 | small | 2023-08-00 | Brazil_Roraima_Mucajai | BR_2015-2023 |
| PP153988.1 | medium | 2023-08-00 | Brazil_Roraima_Mucajai | BR_2015-2023 |
| PP153989.1 | long | 2023-08-00 | Brazil_Roraima_Mucajai | BR_2015-2023 |
| PP153990.1 | small | 2023-08-00 | Brazil_Roraima_Mucajai | BR_2015-2023 |
| PP153991.1 | medium | 2023-08-00 | Brazil_Roraima_Mucajai | BR_2015-2023 |
| PP153992.1 | long | 2023-08-00 | Brazil_Roraima_Mucajai | BR_2015-2023 |

| | | | | |
|---|---|---|---|---|
| PP154119.1 | small | 2023-11-00 | Brazil_Amazonas_Manaus | BR_2015-2023 |
| PP154120.1 | medium | 2023-11-00 | Brazil_Amazonas_Manaus | BR_2015-2023 |
| PP154121.1 | long | 2023-11-00 | Brazil_Amazonas_Manaus | BR_2015-2023 |
| PP154122.1 | small | 2023-11-00 | Brazil_Amazonas_Manaus | BR_2015-2023 |
| PP154123.1 | medium | 2023-11-00 | Brazil_Amazonas_Manaus | BR_2015-2023 |
| PP154124.1 | long | 2023-11-00 | Brazil_Amazonas_Manaus | BR_2015-2023 |
| PP154125.1 | small | 2023-11-00 | Brazil_Amazonas_Manaus | BR_2015-2023 |
| PP154126.1 | medium | 2023-11-00 | Brazil_Amazonas_Manaus | BR_2015-2023 |
| PP154127.1 | long | 2023-11-00 | Brazil_Amazonas_Manaus | BR_2015-2023 |
| PP154041.1 | small | 2023-12-00 | Brazil_Amazonas_Manaus | BR_2015-2023 |
| PP154042.1 | medium | 2023-12-00 | Brazil_Amazonas_Manaus | BR_2015-2023 |
| PP154043.1 | long | 2023-12-00 | Brazil_Amazonas_Manaus | BR_2015-2023 |
| PP154044.1 | small | 2023-12-00 | Brazil_Amazonas_Manaus | BR_2015-2023 |
| PP154045.1 | medium | 2023-12-00 | Brazil_Amazonas_Manaus | BR_2015-2023 |
| PP154046.1 | long | 2023-12-00 | Brazil_Amazonas_Manaus | BR_2015-2023 |
| PP154047.1 | small | 2023-12-00 | Brazil_Amazonas_Manaus | BR_2015-2023 |
| PP154048.1 | medium | 2023-12-00 | Brazil_Amazonas_Manaus | BR_2015-2023 |
| PP154049.1 | long | 2023-12-00 | Brazil_Amazonas_Manaus | BR_2015-2023 |
| PP154050.1 | small | 2023-12-00 | Brazil_Amazonas_Manaus | BR_2015-2023 |
| PP154051.1 | medium | 2023-12-00 | Brazil_Amazonas_Manaus | BR_2015-2023 |
| PP154052.1 | long | 2023-12-00 | Brazil_Amazonas_Manaus | BR_2015-2023 |
| PP154053.1 | small | 2023-12-00 | Brazil_Amazonas_Manaus | BR_2015-2023 |
| PP154054.1 | medium | 2023-12-00 | Brazil_Amazonas_Manaus | BR_2015-2023 |
| PP154055.1 | long | 2023-12-00 | Brazil_Amazonas_Manaus | BR_2015-2023 |
| PP154056.1 | small | 2023-12-00 | Brazil_Amazonas_Manaus | BR_2015-2023 |
| PP154057.1 | medium | 2023-12-00 | Brazil_Amazonas_Manaus | BR_2015-2023 |
| PP154058.1 | long | 2023-12-00 | Brazil_Amazonas_Manaus | BR_2015-2023 |
| PP154059.1 | small | 2023-12-00 | Brazil_Amazonas_Manaus | BR_2015-2023 |
| PP154060.1 | medium | 2023-12-00 | Brazil_Amazonas_Manaus | BR_2015-2023 |
| PP154061.1 | long | 2023-12-00 | Brazil_Amazonas_Manaus | BR_2015-2023 |
| PP154062.1 | small | 2023-12-00 | Brazil_Amazonas_Manaus | BR_2015-2023 |
| PP154063.1 | medium | 2023-12-00 | Brazil_Amazonas_Manaus | BR_2015-2023 |
| PP154064.1 | long | 2023-12-00 | Brazil_Amazonas_Manaus | BR_2015-2023 |
| PP154065.1 | small | 2023-12-00 | Brazil_Amazonas_Manaus | BR_2015-2023 |
| PP154066.1 | medium | 2023-12-00 | Brazil_Amazonas_Manaus | BR_2015-2023 |
| PP154067.1 | long | 2023-12-00 | Brazil_Amazonas_Manaus | BR_2015-2023 |
| PP154068.1 | small | 2023-12-00 | Brazil_Amazonas_Manaus | BR_2015-2023 |
| PP154069.1 | medium | 2023-12-00 | Brazil_Amazonas_Manaus | BR_2015-2023 |
| PP154070.1 | long | 2023-12-00 | Brazil_Amazonas_Manaus | BR_2015-2023 |
| PP154071.1 | small | 2023-12-00 | Brazil_Amazonas_Manaus | BR_2015-2023 |
| PP154072.1 | medium | 2023-12-00 | Brazil_Amazonas_Manaus | BR_2015-2023 |
| PP154073.1 | long | 2023-12-00 | Brazil_Amazonas_Manaus | BR_2015-2023 |

| | | | | |
|---|---|---|---|---|
| PP154074.1 | small | 2023-12-00 | Brazil_Amazonas_Manaus | BR_2015-2023 |
| PP154075.1 | medium | 2023-12-00 | Brazil_Amazonas_Manaus | BR_2015-2023 |
| PP154076.1 | long | 2023-12-00 | Brazil_Amazonas_Manaus | BR_2015-2023 |
| PP154077.1 | small | 2023-12-00 | Brazil_Amazonas_Manaus | BR_2015-2023 |
| PP154078.1 | medium | 2023-12-00 | Brazil_Amazonas_Manaus | BR_2015-2023 |
| PP154079.1 | long | 2023-12-00 | Brazil_Amazonas_Manaus | BR_2015-2023 |
| PP154080.1 | small | 2023-12-00 | Brazil_Amazonas_Manaus | BR_2015-2023 |
| PP154081.1 | medium | 2023-12-00 | Brazil_Amazonas_Manaus | BR_2015-2023 |
| PP154082.1 | long | 2023-12-00 | Brazil_Amazonas_Manaus | BR_2015-2023 |
| PP154083.1 | small | 2023-12-00 | Brazil_Amazonas_Manaus | BR_2015-2023 |
| PP154084.1 | medium | 2023-12-00 | Brazil_Amazonas_Manaus | BR_2015-2023 |
| PP154085.1 | long | 2023-12-00 | Brazil_Amazonas_Manaus | BR_2015-2023 |
| PP154086.1 | small | 2023-12-00 | Brazil_Amazonas_Manaus | BR_2015-2023 |
| PP154087.1 | medium | 2023-12-00 | Brazil_Amazonas_Manaus | BR_2015-2023 |
| PP154088.1 | long | 2023-12-00 | Brazil_Amazonas_Manaus | BR_2015-2023 |
| PP154089.1 | small | 2023-12-00 | Brazil_Amazonas_Manaus | BR_2015-2023 |
| PP154090.1 | medium | 2023-12-00 | Brazil_Amazonas_Manaus | BR_2015-2023 |
| PP154091.1 | long | 2023-12-00 | Brazil_Amazonas_Manaus | BR_2015-2023 |
| PP154092.1 | small | 2023-12-00 | Brazil_Amazonas_Manaus | BR_2015-2023 |
| PP154093.1 | medium | 2023-12-00 | Brazil_Amazonas_Manaus | BR_2015-2023 |
| PP154094.1 | long | 2023-12-00 | Brazil_Amazonas_Manaus | BR_2015-2023 |
| PP154095.1 | small | 2023-12-00 | Brazil_Amazonas_Manaus | BR_2015-2023 |
| PP154096.1 | medium | 2023-12-00 | Brazil_Amazonas_Manaus | BR_2015-2023 |
| PP154097.1 | long | 2023-12-00 | Brazil_Amazonas_Manaus | BR_2015-2023 |
| PP154098.1 | small | 2023-12-00 | Brazil_Amazonas_Manaus | BR_2015-2023 |
| PP154099.1 | medium | 2023-12-00 | Brazil_Amazonas_Manaus | BR_2015-2023 |
| PP154100.1 | long | 2023-12-00 | Brazil_Amazonas_Manaus | BR_2015-2023 |
| PP154101.1 | small | 2023-12-00 | Brazil_Amazonas_Manaus | BR_2015-2023 |
| PP154102.1 | medium | 2023-12-00 | Brazil_Amazonas_Manaus | BR_2015-2023 |
| PP154103.1 | long | 2023-12-00 | Brazil_Amazonas_Manaus | BR_2015-2023 |
| PP154104.1 | small | 2023-12-00 | Brazil_Amazonas_Manaus | BR_2015-2023 |
| PP154105.1 | medium | 2023-12-00 | Brazil_Amazonas_Manaus | BR_2015-2023 |
| PP154106.1 | long | 2023-12-00 | Brazil_Amazonas_Manaus | BR_2015-2023 |
| PP154107.1 | small | 2023-12-00 | Brazil_Amazonas_Manaus | BR_2015-2023 |
| PP154108.1 | medium | 2023-12-00 | Brazil_Amazonas_Manaus | BR_2015-2023 |
| PP154109.1 | long | 2023-12-00 | Brazil_Amazonas_Manaus | BR_2015-2023 |
| PP154110.1 | small | 2023-12-00 | Brazil_Amazonas_Manaus | BR_2015-2023 |
| PP154111.1 | medium | 2023-12-00 | Brazil_Amazonas_Manaus | BR_2015-2023 |
| PP154112.1 | long | 2023-12-00 | Brazil_Amazonas_Manaus | BR_2015-2023 |
| PP154113.1 | small | 2023-12-00 | Brazil_Amazonas_Manaus | BR_2015-2023 |
| PP154114.1 | medium | 2023-12-00 | Brazil_Amazonas_Manaus | BR_2015-2023 |
| PP154115.1 | long | 2023-12-00 | Brazil_Amazonas_Manaus | BR_2015-2023 |

| | | | | |
|---|---|---|---|---|
| PP154116.1 | small | 2023-12-00 | Brazil_Amazonas_Manaus | BR_2015-2023 |
| PP154117.1 | medium | 2023-12-00 | Brazil_Amazonas_Manaus | BR_2015-2023 |
| PP154118.1 | long | 2023-12-00 | Brazil_Amazonas_Manaus | BR_2015-2023 |
| LEIAL2076_ES01_M | medium | 2024-04-00 | Brazil_EspiritoSanto_Colatina | ES_2024 |
| LEIAL2076_ES01_S | small | 2024-04-00 | Brazil_EspiritoSanto_Colatina | ES_2024 |
| LEIAL2077_ES02_M | medium | 2024-04-00 | Brazil_EspiritoSanto_Colatina | ES_2024 |
| LEIAL2077_ES02_S | small | 2024-04-00 | Brazil_EspiritoSanto_Colatina | ES_2024 |
| LEIAL2078_ES03_M | medium | 2024-04-00 | Brazil_EspiritoSanto_RioBananal | ES_2024 |
| LEIAL2078_ES03_S | small | 2024-04-00 | Brazil_EspiritoSanto_RioBananal | ES_2024 |
| LEIAL2079_ES04_M | medium | 2024-03-00 | Brazil_EspiritoSanto_RioBananal | ES_2024 |
| LEIAL2079_ES04_S | small | 2024-03-00 | Brazil_EspiritoSanto_RioBananal | ES_2024 |
| LEIAL2080_ES05_M | medium | 2024-04-00 | Brazil_EspiritoSanto_LaranjaTerra | ES_2024 |
| LEIAL2080_ES05_S | small | 2024-04-00 | Brazil_EspiritoSanto_LaranjaTerra | ES_2024 |
| LEIAL2081_ES06_M | medium | 2024-04-00 | Brazil_EspiritoSanto_LaranjaTerra | ES_2024 |
| LEIAL2081_ES06_S | small | 2024-04-00 | Brazil_EspiritoSanto_LaranjaTerra | ES_2024 |
| 321559213_L | long | 2024-04-00 | Brazil_EspiritoSanto_Colatina | ES_2024 |
| 321559213_M | medium | 2024-04-00 | Brazil_EspiritoSanto_Colatina | ES_2024 |
| 321559213_S | small | 2024-04-00 | Brazil_EspiritoSanto_Colatina | ES_2024 |

**Figure S1**. The boxplots compare the Ct values of individuals with and without each symptom. Horizontal bars represent the Ct medians and interquartile ranges (IQR). Two-sided P-values for the nonparametric Mann–Whitney test are shown for each group

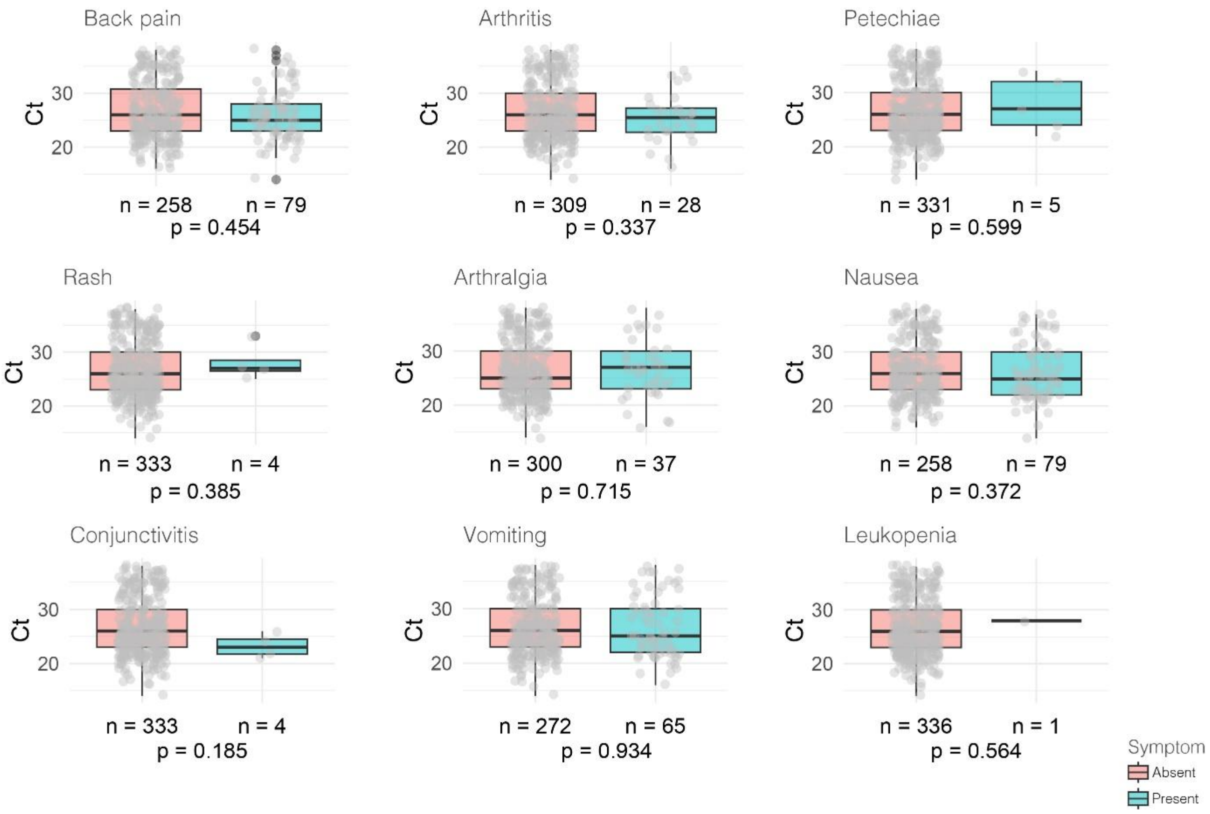
